# The Role of Tumor Necrosis Factor Signaling in Atherosclerosis and Stroke

**DOI:** 10.64898/2026.04.29.26352088

**Authors:** Jane Buckley, Stephen Brennan, Katie Harris, Eoin Brennan, Pol Camps-Renon, Tim Cassidy, Sarah Gorey, Pablo Hervella, Ramon Iglesias-Rey, Gordon Lowe, Jonathan McNulty, Fawad Ahmad, Francisco Purroy, Mikel Vicente-Pascual, Dylan G. Ryan, Padraig Synnott, Cathal Walsh, Paul Welsh, David Williams, Mark Woodward, Peter J. Kelly, John J. McCabe

**Affiliations:** Health Research Board (HRB) Stroke Clinical Trials Network Ireland (SCTNI), Ireland; School of Medicine, University College Dublin (UCD), Ireland; Stroke Service, Mater Misericordiae University Hospital, Dublin, Ireland; School of Medicine, University of Galway, Ireland; National Institute for Prevention and Cardiovascular Health, Galway, Ireland; The George Institute for Global Health, University of New South Wales, Sydney, Australia; Department of Neurology, Institute of Biomedical Research Sant Pau (IIB-Sant Pau), Hospital de la Santa Creu i Sant Pau, Barcelona, Spain; St Vincent’s University Hospital, Dublin, Ireland; Neuroimaging and Biotechnology Laboratory (NOBEL), Health Research Institute of Santiago de Compostela, Spain; School of Cardiovascular and Metabolic Health, University of Glasgow, Glasgow, UK; Department of Neurology, Hospital Universitari Arnau de Vilanova, Lleida, Spain; Department of Clinical Neurosciences, University of Lleida, Spain; Trinity Biomedical Sciences Institute, Trinity College Dublin, Ireland; Department of Biostatistics, Trinity College Dublin, Ireland; Department of Geriatric and Stroke Medicine, RCSI University of Medicine and Health Sciences Dublin, Leinster, Ireland; Department of Geriatric and Stroke Medicine, Beaumont Hospital, Dublin, Ireland; The George Institute for Global Health, School of Public Health, Imperial College London, London, UK; Nuffield Department of Women’s and Reproductive Health, University of Oxford, Oxford, UK

**Keywords:** Stroke, tumor necrosis factor, prognosis, recurrence, atherosclerosis, prevention

## Abstract

**Background and Objectives:** Inflammation is an emerging target for stroke prevention, but additional therapeutic candidates are needed. Experimental and observational studies implicate tumor necrosis factor (TNF) in atherosclerotic plaque progression and cardiovascular events. We integrated plasma proteomics from a population-based study and prospective stroke cohorts together with transcriptomic profiling of atherosclerotic plaque to investigate TNF signaling in stroke pathogenesis.

**Methods:** First, we assessed associations between 34 TNF-Superfamily proteins (Olink) and risk of incident ischemic stroke, myocardial infarction (MI), and peripheral vascular disease (PVD) among 45,783 participants from a population-based study (UK Biobank) without cardiovascular disease or autoimmune disease at baseline. Second, we performed an individual-participant data (IPD) meta-analysis of four prospective cohorts (n=2,051), identified by systematic review, to examine associations between TNF-α and risk of vascular recurrence after ischemic stroke. Third, we characterised plaque-level TNF-biology using bulk transcriptomic data from 43 plaque segments (24 patients) and Xenium spatial transcriptomics (12 donors).

**Results:** In UK Biobank, after multivariable adjustment for cardiovascular risk factors, circulating TNF-α, TNFR1 and TNFR2 were associated with incident stroke (Hazard Ratio per SD increase, 1.15, [95% CI 1.08–1.23], 1.21, [1.13–1.30], and 1.21 [1.13–1.30], respectively) with similar findings for MI and PVD. 18 additional Superfamily members were associated with incident stroke. In the IPD analysis of stroke cohorts, TNF-α was independently associated with recurrent stroke (Risk Ratio (RR) 1.47, 1.06–2.03; top vs. bottom third) and major adverse cardiovascular events (MACE) (RR 1.49, 1.09–2.04) after adjustment for cardiovascular risk factors and secondary prevention medication use (537 MACE events, 6290 person-years follow up). In transcriptomic analyses, a TNF-axis score was higher in unstable than stable carotid plaque segments (difference 1.47 SD, 95% CI 1.02-1.92). Spatial transcriptomics showed that TNF was more frequently detected in the fibrous cap than in the media (OR 2.29, 95% CI 1.88-2.77).

**Discussion:** TNF signaling is independently associated with incident ischemic stroke and recurrent stroke/MACE after stroke. Supporting evidence from human carotid plaque transcriptomics also implicate TNF signalling in atherosclerosis. These results support evaluation of TNF-targeted therapies for stroke prevention.

## Introduction

Five-year vascular recurrence rates after stroke are as high as 20-30% and have stagnated despite widespread use of guideline-based secondary prevention.^1^ Several lines of evidence implicate inflammatory mechanisms in vascular recurrence after stroke^2-5^ and inflammation is a promising target for prevention of stroke and cardiovascular disease.^6-8^ Randomized controlled trials (RCTs) demonstrated proof of concept that anti-inflammatory therapies targeting the NLRP3)/Interleukin-1 beta (IL-1β)/Interleukin-6 (IL-6) signaling pathway with colchicine and canakinumab can reduce the risk of major adverse cardiovascular events (MACE) in patients with coronary artery disease (CAD).^9-12^ Several RCTs evaluating direct IL-6 inhibition for vascular prevention are underway or completed.^13^ However, alternative immune signaling pathways are also implicated in atherosclerosis, but have not been prioritised for targeted therapies.

The Tumor Necrosis Factor superfamily (TNFSF) constitutes a complex signaling regulatory system comprising of multiple ligands and receptors. Of the TNFSF, TNF-α plays a dominant role due to the ubiquitous nature of its primary receptors TNFR1 and TNFR2^14^ - through which the resultant signaling cascade, activates NF-κB-mediated inflammatory pathways that regulate leukocyte recruitment, endothelial activation, and matrix remodeling. Multiple lines of evidence implicate TNF-α and the TNFSF in cardiovascular disease. Experimental studies have established the central role of TNF signaling in the development of atherosclerosis^15-17^ and circulating TNF-α levels are independently associated with recurrent MACE in patients with CAD.^18^ Observational studies of TNF-driven diseases such as Rheumatoid Arthritis (RA) demonstrate a 1.5-2 fold increased risk of incident cardiovascular events and stroke^19^ – a risk markedly attenuated when TNF inhibitors are utilized in RA disease management.^20^ However, TNF inhibitors have not been evaluated in RCTs for prevention of stroke and MACE.

We used multiple lines of evidence to investigate the role of TNF, TNF Receptors and the wider TNFSF in incident (first) cardiovascular events and recurrent vascular events after stroke. First, we used proteomic data from a population-based study to investigate the association between TNF-α/TNFSF with incident stroke, CAD and peripheral vascular disease (PVD) in participants without a history of vascular disease. Second, we used individual participant data (IPD) from four prospective studies to explore the association between TNF-α measurements after ischemic stroke and risk of recurrent stroke and MACE. Finally, because circulating levels cannot localize TNF signaling to the vessel wall, we used bulk and spatial transcriptomics to examine TNF expression in human carotid atherosclerotic plaques.

## Methods

### TNF Signaling and Incident Stroke and Related Cardiovascular Endpoints

We leveraged proteomic data from 54,219 participants from the UK Biobank, a large, population-based prospective cohort study.^21^ Plasma levels of 34 TNFSF proteins, including 12 ligands, 20 receptors and 2 induced intracellular proteins, were measured using Olink Explore. We excluded participants with prevalent cardiovascular disease, atrial fibrillation (AF), end-stage renal disease (ESRD), and self-reported rheumatological or autoimmune disease (Table S1) at baseline.

Protein abundances were adjusted for assay plate using protein-specific random-intercept linear mixed models, and the resulting standardized residuals were entered into Cox proportional hazards models to assess the association of each TNFSF member with incident ischemic stroke, with cumulative incidence estimated using the Kaplan-Meier method. We repeated our analysis for myocardial infarction (MI) and PVD. Outcomes in UK Biobank were derived from linked hospital admissions data/national death registry records. Models were adjusted for demographic factors, socioeconomic status, and cardiovascular risk factors (age, sex, ethnicity, education, deprivation, smoking, alcohol intake, diabetes, BMI, and systolic blood pressure). We applied the Benjamini-Hochberg method to control the false discovery rate (FDR) across all protein–outcome associations. Sensitivity analyses included: (1) additional adjustment for log-transformed circulating CRP and IL-6; (2) a lagged analysis in which events and person-time accrued during the first 2 years of follow-up were excluded to reduce potential reverse causation; (3) exclusion of participants with baseline CRP >10mg/L to exclude confounding by acute inflammation; and (4) estimation of hazard ratios separately for 0-5, 5-10 and >10 years of follow-up.

### TNF-α and Risk of Recurrent Vascular Events After Stroke

We performed a systematic review and meta-analysis of studies investigating the association between circulating TNF-α, measured after ischemic stroke or transient ischemic attack (TIA), and the risk of recurrent stroke and MACE. Studies contributed data as part of the Blood Inflammatory markers in Stroke Collaboration (BISC) – the methodology of which is described in previous publications.^2^ The review was conducted according to PRISMA Guidelines^22^ and the protocol was registered in the International Prospective Register of Systematic Reviews database (PROSPERO ID: CRD42024619328). A systematic literature search of OVID Medline and EMBASE (1945 to 24^th^ November 2024) for English-language articles featuring the keywords ‘stroke’ and ‘tumor necrosis factor alpha’ and ‘recurrence’. Next, authors of eligible studies were invited to provide IPD for a two-stage meta-analysis. First, within-study Cox regression analysis was performed for all studies with time to event data to estimate the hazard ratio (HR). For the single study with a nested case-control design (PROGRESS), the odds ratio (OR) for recurrent vascular events was estimated by conditional logistic regression. Since outcomes were infrequent, HRs and ORs were assumed to approximate the same relative risk, and results are collectively summarised as risk ratios (RR). We then performed pooled meta-analysis of all studies using random effects meta-analysis with inverse variance weighting. TNF-α measurements were log_e_-transformed to stabilise the variance. Analyses were conducted per unit and per third increase log_e_-TNF-α with increasing degrees of adjustment. Model 1 adjusted for age, sex, index event, hypertension, smoking, diabetes mellitus, CAD, AF, statin use, antithrombotic therapy and trial arm (where appropriate). Models 2 and 3 adjusted for the variables in Model 1 and loge-IL-6 and loge-high-sensitivity C-Reactive Protein (hsCRP), respectively. Only outcome events occurring after sample measurement were included. We performed several sensitivity analyses: 1) restricted to the two largest studies; 2) excluding the study without time-to-event data, and 3) a leave-one-out analysis to explore the influence of a single-study on the results. Subgroup analyses were performed by timing of phlebotomy and study population (TIA only vs. other/mixed). Between-group differences were assessed using a test for interaction.

Further details regarding the search strategy, eligibility criteria, and data harmonisation procedures are provided in Supplemental Methods. One study (PROGRESS) could not directly share IPD for centralised pooling and for this study a pre-specified identical analysis was performed locally with summary aggregate data provided for two-stage meta-analysis with the other studies.

### Carotid Plaque Transcriptomic Analyses

To examine TNF signaling within atherosclerotic plaques, we analyzed two publicly available datasets from carotid endarterectomy specimens: (i) bulk transcriptomic data from 43 histologically classified stable and unstable plaque segments across 24 patients,^23^ and (ii) Xenium (10x Genomics) spatial transcriptomic data from 12 with pathologist-annotated subregions from 12 donors.^24^

For the bulk transcriptomic analysis, we used data from the Maastricht Human Plaque Study (GEO accession GSE163154; Illumina HumanRef-8 v2), which included 43 plaque segments from 24 symptomatic men.^23^ Plaque segments were histologically classified by pathologists as stable (fibrous cap atheroma or intimal thickening; n=16) or unstable (intraplaque hemorrhage, rupture or occlusion; n=27). We compared a TNF-axis score (mean standardized expression of TNF, TNFR1 and TNFR2), TNF expression and TNF-α/NF-κB pathway scores (Table S2) between unstable and stable plaque segments using linear mixed-effects models with a random intercept for patient. Pathway scores were calculated using single-sample gene set enrichment analysis (ssGSEA) of the Hallmark TNF-α signaling via NF-κB gene set, excluding TNF to avoid overlap with the primary TNF-axis score. Gene-level differential expression and gene set enrichment were assessed using limma and cameraPR.

For the spatial transcriptomic analysis, TNF detection was defined as ≥1 raw transcript per cell. We compared TNF detection across plaque regions (fibrous cap, necrotic core, intima and media) using mixed-effects logistic regression, with media as the reference category and a random intercept for each donor. Sensitivity analyses used patient-level binomial models and a ≥2-transcript detection threshold.

### Ethics Approval / Data Access

UK Biobank has ethical approval from the North West Multi-Centre Research Ethics Committee (REC reference 11/NW/03820). Participants provided written informed consent. UK Biobank analyses were conducted under project approval 1282644 and in accordance with the Declaration of Helsinki. This study adheres to the STROBE reporting guidelines.^25^

## Results

### TNF Signaling and Incident Stroke/Related Cardiovascular Endpoints

Among 45,783 UK Biobank participants free of cardiovascular disease, AF, ESRD and autoimmune disease at baseline, there were 1,509 MIs, 856 ischemic strokes, and 783 PVD events over a median follow-up of 15.2 years (IQI 14.4–15.9). Among participants with incident ischemic stroke, the median time from blood sampling to event was 9.2 years (IQI 5.9–11.9). Baseline demographics are provided in the Table S3. After adjustment for age, sex, ethnicity, deprivation, and vascular risk factors, TNF-α (HR 1.15, 95% CI 1.08–1.23), TNFR1 (HR 1.21, 95% CI 1.13–1.30) and TNFR2 (HR 1.21, 95% CI 1.13–1.30) were each independently associated with incident stroke (analysed per SD increase) (Table S4). Compared with the lowest third, participants in the highest third of TNF-α and its soluble receptors had a 31–47% higher risk of ischemic stroke (TNF-α: HR 1.47, 95% CI 1.21–1.78; TNFR1: HR 1.31, 95% CI 1.07–1.60; TNFR2: HR 1.35, 95% CI 1.11–1.63; all p-trend <0.01) (Figure 1). Of the remaining members of the TNFSF analysed, two ligands (APRIL, CD70) and 16 receptors (TACI, BCMA, RANK, CD27, CD40, LTBR, HVEM, OPG, DR4, DR5, DR6, Fn14, FAS, OX40, RELT, TROY) were also significantly associated with higher risk of incident stroke per SD increase (Figure 1, Table S4). One ligand (FASL) was associated with a lower risk of stroke.

**Figure 1:**
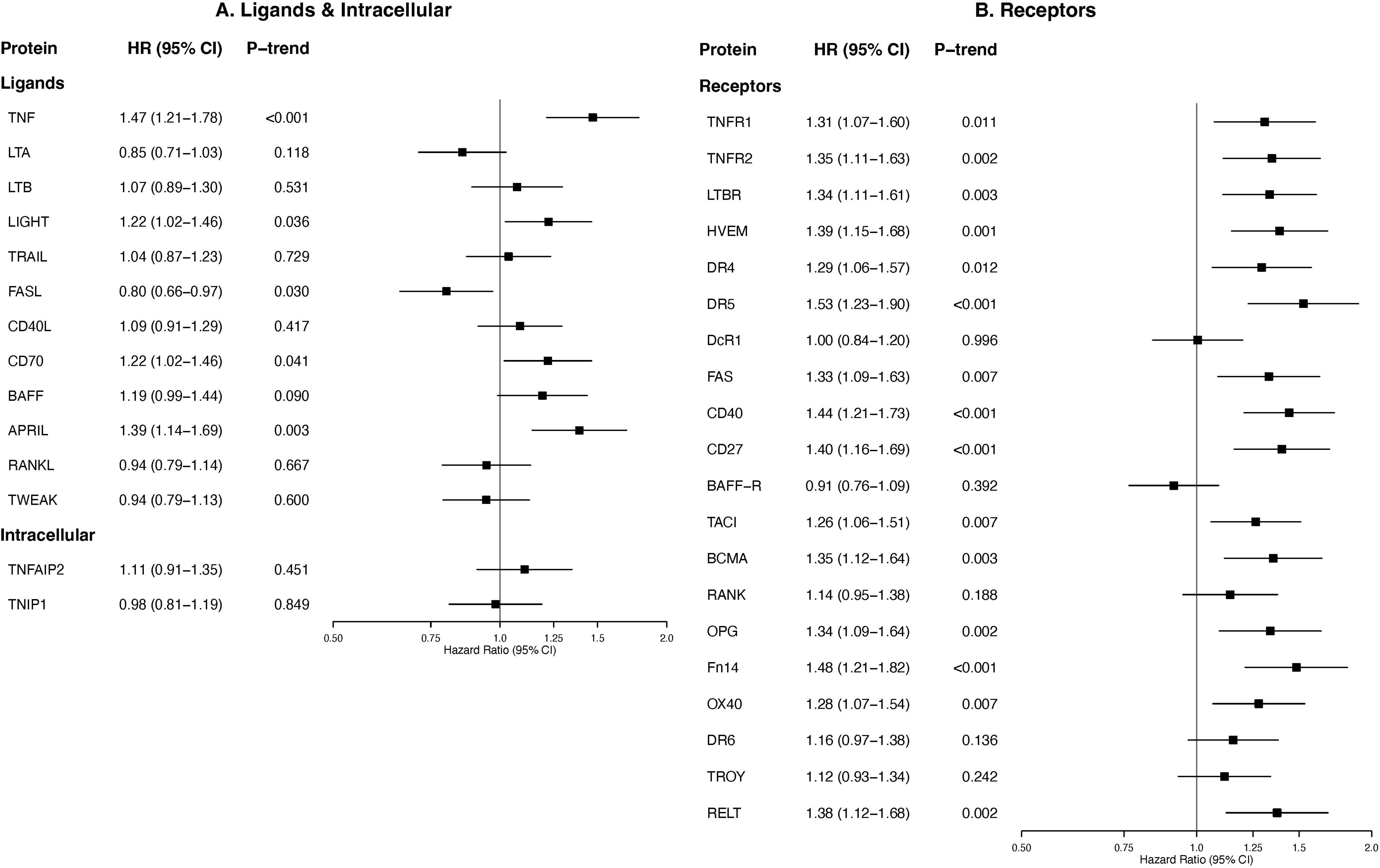
TNF-α, TNF receptors, and TNF Superfamily ligands and Receptors, and Risk of Incident Ischemic stroke. Associations are expressed for top vs bottom third for circulating protein, after adjustment for age, sex, ethnicity and cardiovascular risk factors. P-trend values are FDR- adjusted for multiple comparisons, confidence intervals are unadjusted. Tertile cutoffs used are assay-specific and cannot be translated to clinical decision thresholds currently. Associations per SD increase are given in Table S4.

Associations were similar for MI and PVD. Higher circulating TNF-α levels were associated with MI (T3 vs T1: HR 1.38, 95% CI 1.20–1.59) and PVD (HR 1.56, 95% CI 1.28–1.90). Similar findings were observed for TNFR1 and TNFR2 (Figures 2-3). Cumulative incidence of each outcome was highest in the top third of circulating TNF, TNFR1 and TNFR2, with separation persisting throughout follow-up (Figures S1–S3). Effect estimates for all outcomes were largely unchanged after additional adjustment for CRP and IL-6 (Figure S4, Table S5), after exclusion of the first two years of follow-up (Figure S5, Table S6) or of participants with baseline CRP >10 mg/L (Figure S6, Table S7). Associations were consistent across 0–5, 5–10 and >10 years of follow-up (Figure S7, Table S8).

**Figure 2:**
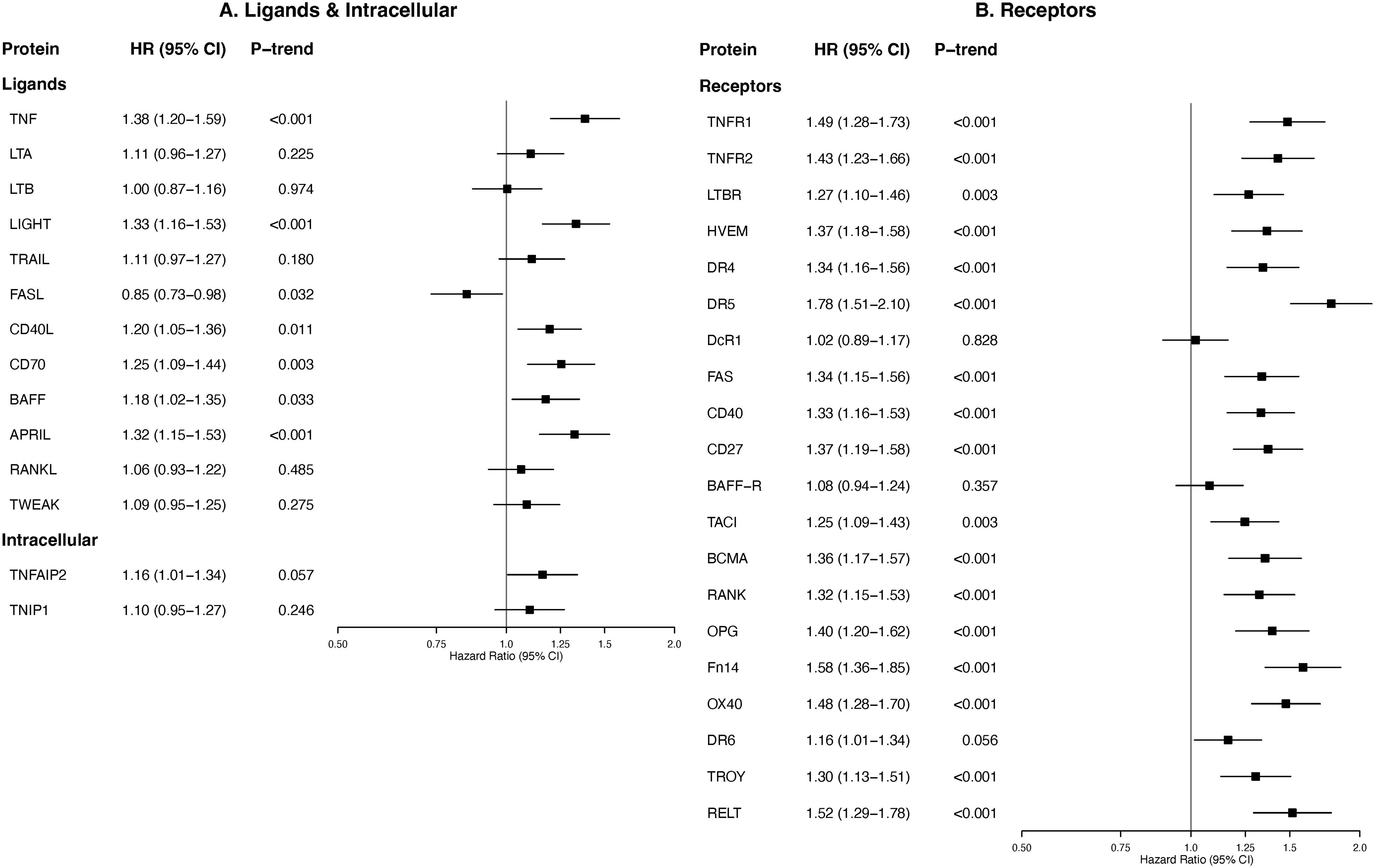
TNF-α, TNF receptors, and TNF Superfamily ligands and Receptors, and Risk of Incident Myocardial Infarction. Associations are expressed for top vs bottom third for circulating protein, after adjustment for age, sex, ethnicity and cardiovascular risk factors. P-trend values are FDR-adjusted for multiple comparisons, confidence intervals are unadjusted. Tertile cutoffs used are assay-specific and cannot be translated to clinical decision thresholds currently. Associations per SD increase are given in Table S4.

**Figure 3:**
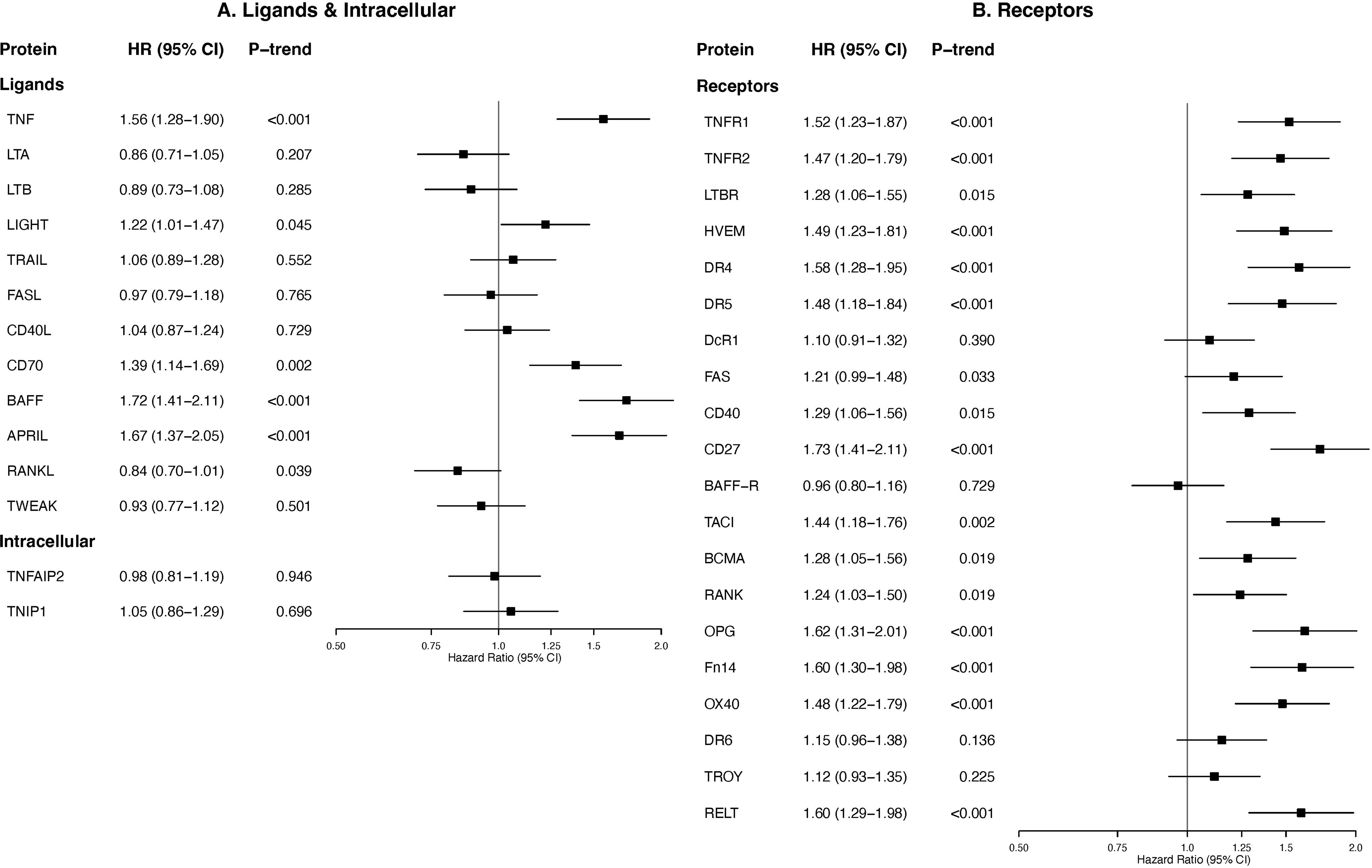
TNF-α, TNF receptors, and TNF Superfamily ligands and Receptors, and Risk of Incident Peripheral Vascular Disease. Associations are expressed for top vs bottom third for circulating protein, after adjustment for age, sex, ethnicity and cardiovascular risk factors. P-trend values are FDR-adjusted for multiple comparisons, confidence intervals are unadjusted. Tertile cutoffs used are assay-specific and cannot be translated to clinical decision thresholds currently.

### TNF-α and Recurrent Vascular Events after Stroke: Systematic Review and IPD Meta-analysis

The systematic review search returned 1168 records, of which 37 studies were selected for full text review (Figure S8). Four eligible studies were identified, including three prospective observational cohort studies and one nested case-control study within an RCT (PROGRESS).^26-29^ 2,051 patients were included in the meta-analysis with 6290 patient-years of follow up. The baseline characteristics are summarised in Table 1. The qualifying index event was ischemic stroke in 1422 (69.3%) and TIA in 629 patients (30.6%). Three studies performed phlebotomy in the acute phase (within 14 days of index event) after the index event, and one study obtained samples in the convalescent phase (median 300 days, IQI 111-753). Additional information regarding the design, methodology, and stroke aetiology of each contributing study is provided in the Supplemental Results and Tables S9-11. The Quality in Prognostic Studies (QUIPS) risk of bias tool demonstrated studies were at low risk of bias (Table S12).

**Table 1:**
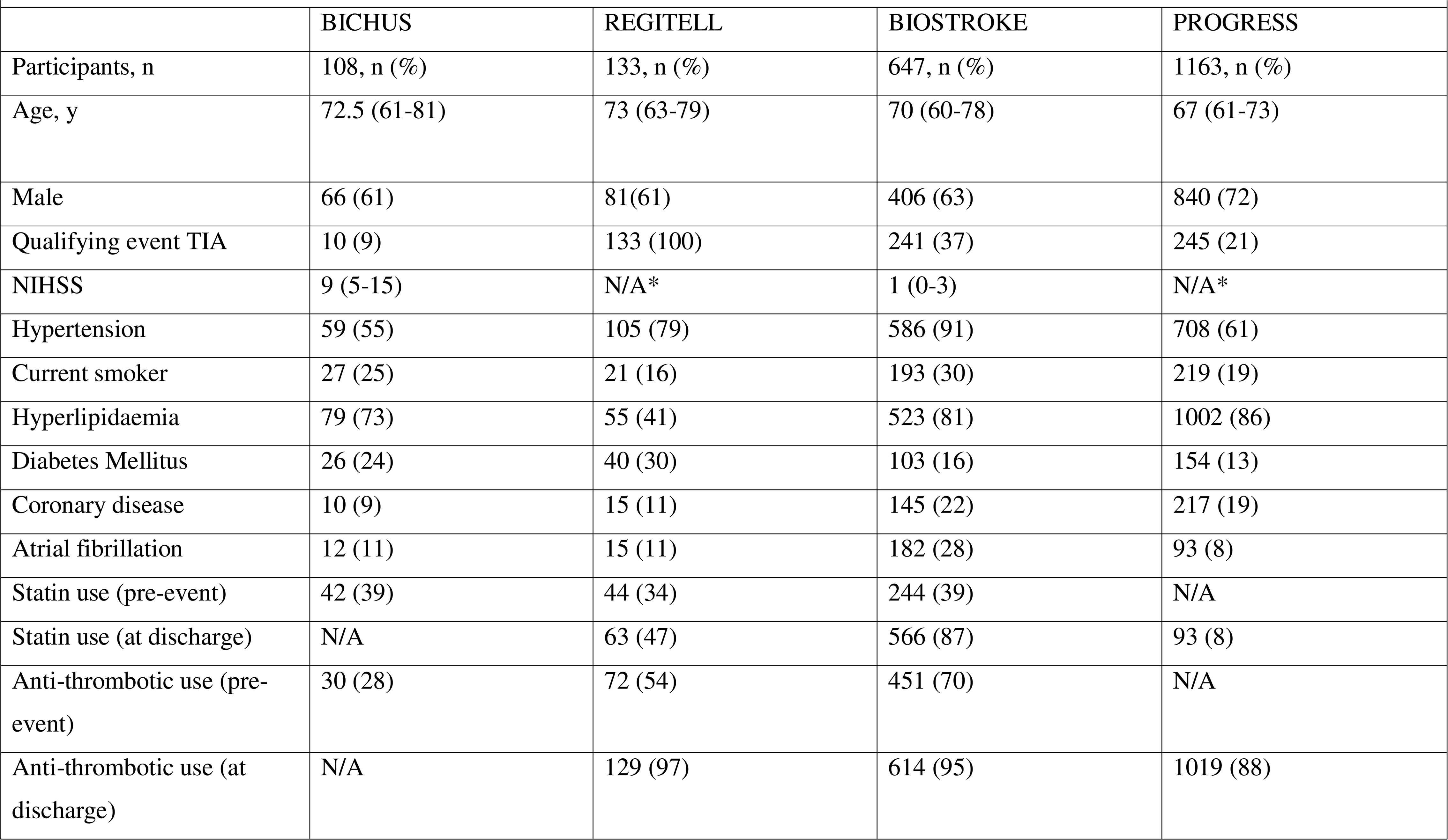
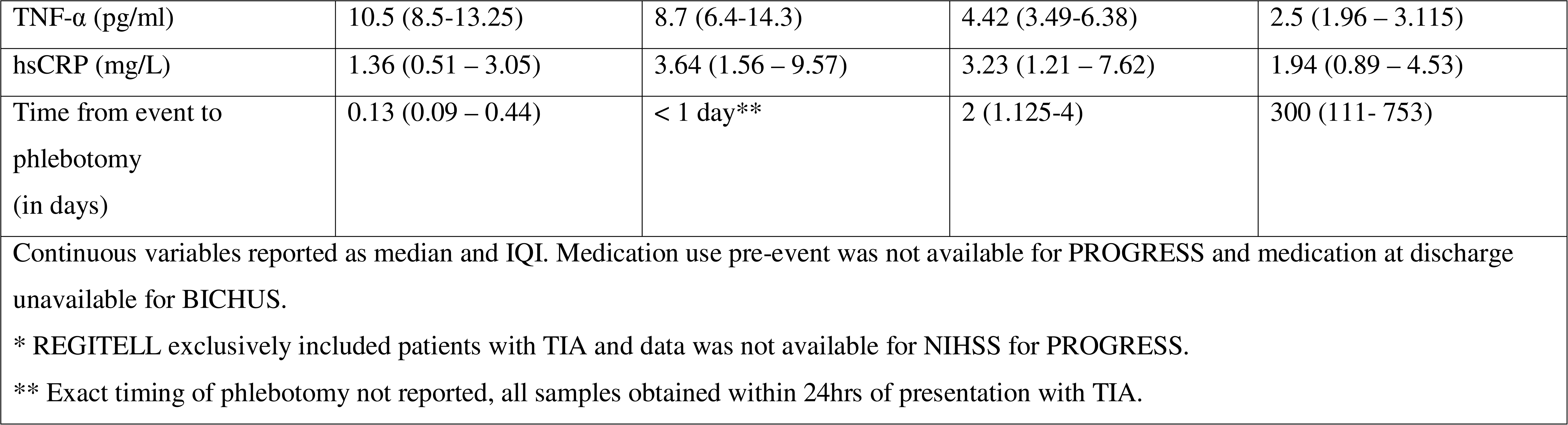
Baseline Characteristics of Participants of IPD Meta-analysis Stratified by Study.

|  | BICHUS | REGITELL | BIOSTROKE | PROGRESS |
|---|---|---|---|---|
| Participants, n | 108, n (%) | 133, n (%) | 647, n (%) | 1163, n (%) |
| Age, y | 72.5 (61-81) | 73 (63-79) | 70 (60-78) | 67 (61-73) |
| Male | 66 (61) | 81(61) | 406 (63) | 840 (72) |
| Qualifying event TIA | 10 (9) | 133 (100) | 241 (37) | 245 (21) |
| NIHSS | 9 (5-15) | N/A* | 1 (0-3) | N/A* |
| Hypertension | 59 (55) | 105 (79) | 586 (91) | 708 (61) |
| Current smoker | 27 (25) | 21 (16) | 193 (30) | 219 (19) |
| Hyperlipidaemia | 79 (73) | 55 (41) | 523 (81) | 1002 (86) |
| Diabetes Mellitus | 26 (24) | 40 (30) | 103 (16) | 154 (13) |
| Coronary disease | 10 (9) | 15 (11) | 145 (22) | 217 (19) |
| Atrial fibrillation | 12 (11) | 15 (11) | 182 (28) | 93 (8) |
| Statin use (pre-event) | 42 (39) | 44 (34) | 244 (39) | N/A |
| Statin use (at discharge) | N/A | 63 (47) | 566 (87) | 93 (8) |
| Anti-thrombotic use (pre-event) | 30 (28) | 72 (54) | 451 (70) | N/A |
| Anti-thrombotic use (at discharge) | N/A | 129 (97) | 614 (95) | 1019 (88) |
| TNF-α (pg/ml) | 10.5 (8.5-13.25) | 8.7 (6.4-14.3) | 4.42 (3.49-6.38) | 2.5 (1.96 – 3.115) |
| hsCRP (mg/L) | 1.36 (0.51 – 3.05) | 3.64 (1.56 – 9.57) | 3.23 (1.21 – 7.62) | 1.94 (0.89 – 4.53) |
| Time from event to phlebotomy<br>(in days) | 0.13 (0.09 – 0.44) | < 1 day** | 2 (1.125-4) | 300 (111- 753) |

Continuous variables reported as median and IQI. Medication use pre-event was not available for PROGRESS and medication at discharge unavailable for BICHUS.
\* REGITELL exclusively included patients with TIA and data was not available for NIHSS for PROGRESS.
\*\* Exact timing of phlebotomy not reported, all samples obtained within 24hrs of presentation with TIA.

In unadjusted analysis, TNF-α levels measured after ischemic stroke/TIA were associated with an increased risk of recurrent stroke during follow-up (pooled RR 1.34, 95% CI 1.05-1.70, per log_e_-unit increase). This association persisted after adjustment for vascular risk factors and medication use (RR 1.36, 95% CI 1.06-1.76). Participants in the highest third of TNF-α levels were observed to have a 50% increased risk of recurrent stroke compared to the lowest third (RR 1.51, 95% CI 1.12 – 2.05), which was not attenuated after adjustment (RR 1.47, 95% CI 1.06-2.03) (Figure 4, Table 2). These associations persisted after mutual adjustment for IL-6 and hsCRP (Table 2).

**Figure 4:**
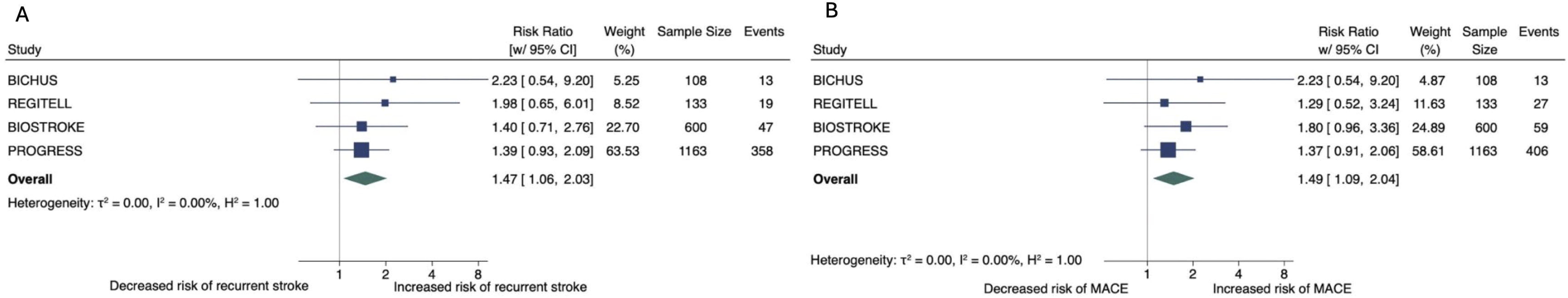
TNF-α and Risk of Recurrent Stroke (A) and Recurrent MACE (B). Associations are expressed for top vs bottom third for circulating protein, after adjustment as per Model 2 (previously described).

**Table 2:**
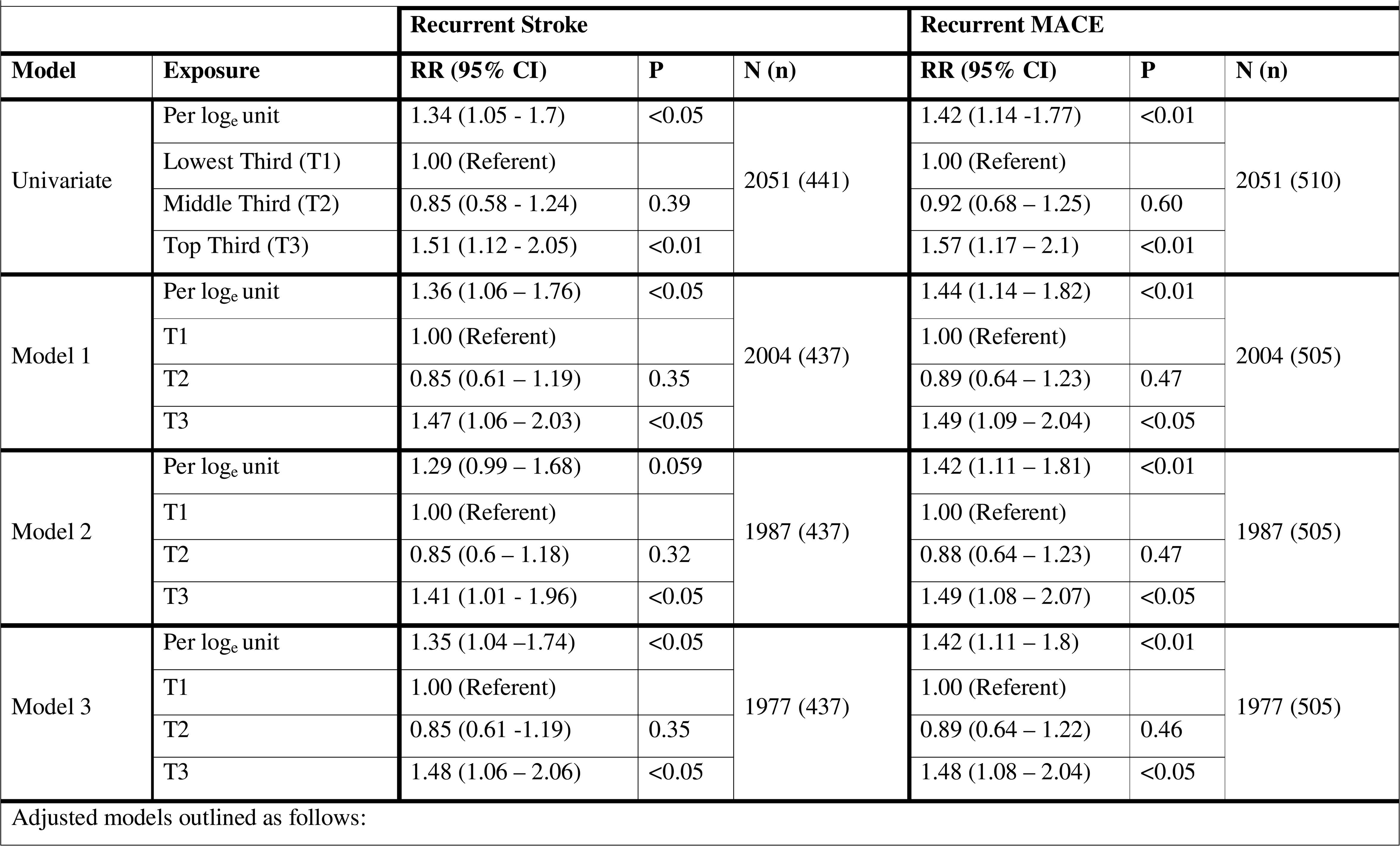
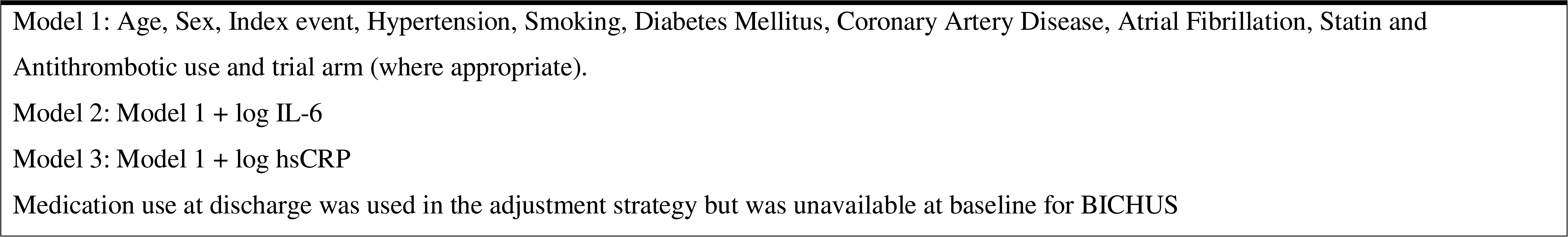
Association between TNF-α and Recurrent Stroke and MACE.

We observed similar results for the secondary outcome of recurrent MACE, with independent associations after adjustment for vascular risk factors and medication use (RR 1.44, 95% CI 1.14-1.82). Those patients in the top third of TNF-α levels had an approximate 50% relative risk increase of MACE compared to the lowest third (RR 1.49, 95% CI 1.09-2.04) (Table 2, Figure 4). These associations remained after additional adjustment for IL-6 or hsCRP.

Associations were largely unchanged across several subgroup and sensitivity analyses. First, TNF-α associations with recurrent stroke and MACE were not modified by timing of measurement after the index event (P_interaction_=0.55 for recurrent stroke, P_interaction_ =0.33 for MACE). Comparing the extreme thirds, TNF-α was associated with recurrent stroke (RR 1.50, 95% CI 1.01-2.24) and MACE (RR 1.62, 95% CI 1.16-2.26) if measured in the acute phase (<14 days). Associations were with directionally-similar, but not statistically significant, in the convalescent window (Table S13). Second, findings remained significant in sensitivity analyses constructed to investigate whether our findings might have been unduly influenced by a single cohort - after excluding the study without time-to-event data (Table S14), on a leave-one-out analysis removing one study at a time (Figure S9), and after restricting the meta-analysis to the two largest studies (Table S15). Third, a subgroup analysis by index event subtype did not show evidence of interaction for recurrent stroke (P_interaction_ 0.74) or MACE (P_interaction_ 0.65) (Table S16).

### TNF signaling in human atherosclerotic plaques

Among 43 carotid plaque segments from 24 patients, the TNF-axis score was higher in unstable than stable segments (difference 1.47 SD, 95% CI 1.02–1.92; P<0.001; Figure 5A). Results were similar in the 16 patients that contributed paired stable and unstable segments (1.47 SD, 95% CI 0.81–2.14; P<0.001) and in analyses accounting for patient clustering (1.47 SD, 95% CI 0.87–2.07; P<0.001). Each of the genes contained in the TNF-axis score (TNF, TNFR1 and TNFR2) was more highly expressed in unstable segments, as was the Hallmark TNF-α signaling via NF-κB pathway score (difference 1.39 SD, 95% CI 0.91–1.87; P<0.001). Gene set enrichment analysis provided further evidence of increased TNF-α/NF-κB signaling in unstable segments (Figure 5B).

**Figure 5.**
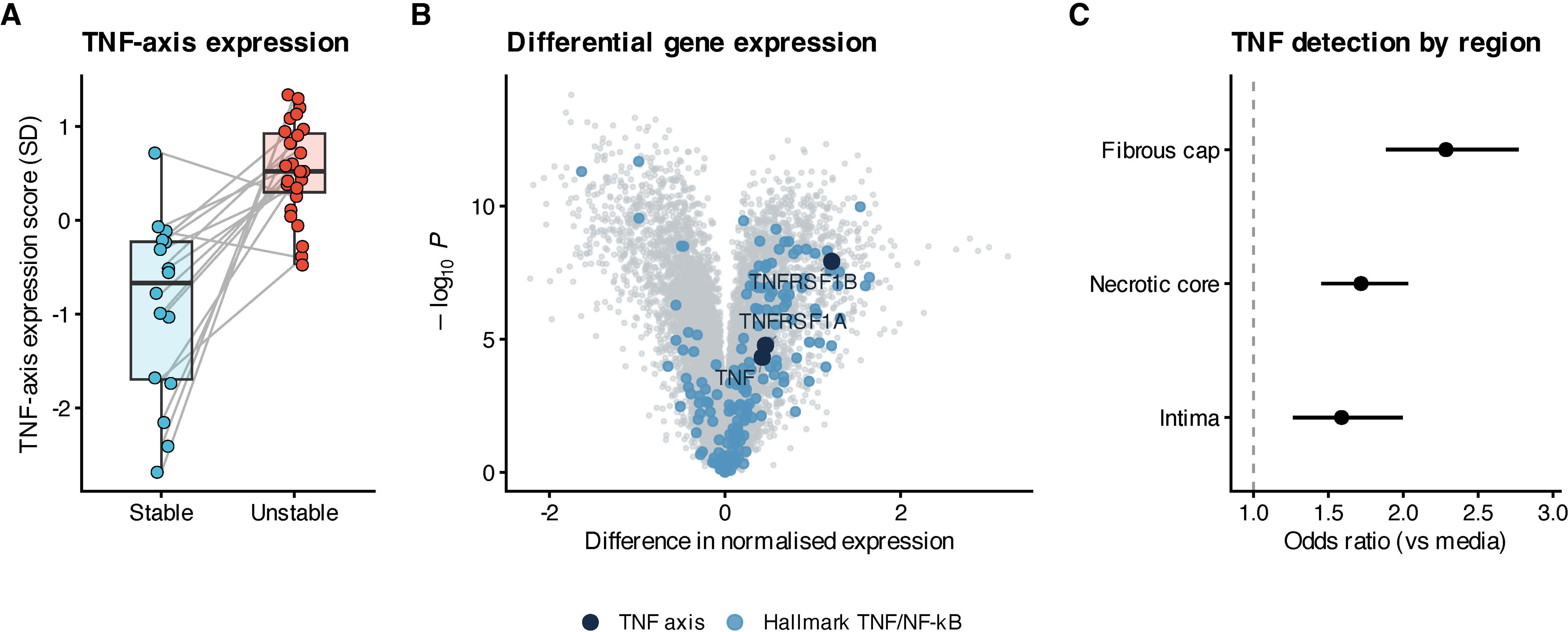
TNF signaling in human carotid atherosclerotic plaques. (A) TNF-axis expression scores in histologically stable (n=16) and unstable (n=27) plaque segments from 24 patients. Lines connect paired segments from the same patient. (B) Differential gene expression between unstable and stable plaques, with TNF-axis and Hallmark TNF-α/NF- κB pathway genes highlighted. (C) TNF detection across plaque regions, reported as odds ratios (95% CI) relative to media.

We next examined TNF expression across different regions of carotid atherosclerotic plaques using spatial transcriptomics. Compared with media, TNF detection was higher in intima (OR 1.59, 95% CI 1.26–2.00), necrotic core (OR 1.72, 95% CI 1.45–2.03), and fibrous cap (OR 2.29, 95% CI 1.88–2.77) (Figure 5C). Findings were consistent in patient-level sensitivity analyses, with greater fibrous cap enrichment when a ≥2-transcript detection threshold was used (OR 6.98, 95% CI 3.08–15.80). At the fibrous cap, macrophages accounted for 62.0% of TNF-positive cells and CD8+ effector T cells for 16.9% (Table S17). TNF detection was low across macrophage subtypes, with the highest median detection in C1Q+ macrophages (2.1%, IQI 0.7–4.4; Table S18).

## Discussion

Our study provides convergent evidence from multiple sources implicating TNF signaling in atherosclerosis and stroke. In a large population-based cohort of apparently-healthy middle-aged adults free of cardiovascular disease, higher circulating TNF-α, TNF receptors, and TNFSF proteins were associated with risk of first ischemic stroke, MI and PVD. Across four prospective studies of patients with ischemic stroke or TIA, higher TNF-α was also independently associated with recurrent stroke and MACE. In human atherosclerotic plaques, TNF expression was higher in unstable plaques than stable plaques and was enriched in the rupture-prone fibrous cap. Together, these findings support further investigation of TNF-related pathways in atherosclerosis and stroke.

Anti-inflammatory therapies hold promise for secondary prevention in patients with CAD and stroke. However, most RCTs have prioritised immune modulation of the IL-1β-IL-6-CRP axis to reduce plaque inflammation and alternative immune targets warrant consideration.^9-13^ TNF-α is a key cytokine in inflammatory signaling and has been successfully targeted for treatment of immune-mediated diseases. Epidemiological studies of patients with RA demonstrate an increased risk of stroke and cardiovascular disease which is not explained by conventional risk factors – with an approximate doubling in relative risk compared to age-matched controls.^19^ Meta-analyses of large cohort studies indicate that TNF inhibitors are associated with a lower risk of stroke, MI, and MACE when used in the management of autoimmune disorders suggesting that these therapies may have pleiotropic plaque-stabilising effects.^20^ Despite consistent evidence from experimental models implicating both TNF-α and the wider TNFSF in atherosclerotic plaque development, RCTs of TNF-inhibitors for vascular prevention have not yet been performed.

Here, we present several lines of evidence from epidemiological data demonstrating the importance of TNF signaling in stroke and atherosclerosis. First, we identified a significant association between higher circulating levels of TNF-α and its two receptors with a 30-40% increased risk of incident ischemic stroke after adjustment for cardiovascular risk factors. A similar effect was seen for two other circulating ligands and 16 other receptors of the TNFSF superfamily further indicating a broad and consistent role for TNF signaling in stroke. These findings were replicated for the atherosclerotic-driven secondary outcomes of MI and PVD. Next, we sought to explore the relationship between circulating TNF-α and recurrent vascular events when measured after stroke. In our IPD meta-analysis of over 2000 patients, we identified a 40-50% increased risk of recurrent stroke and MACE for those in the highest third of TNF-α levels when compared with patients in the bottom third. With each three-fold increase in TNF-α levels, there was an approximate 35% increased risk of recurrent stroke, and 40% increased risk of recurrent MACE. These findings were robust and minimally attenuated after adjusted for cardiovascular risk factors and secondary medication use and in subgroup analyses accounting for between-study differences in timing of sample measurement and stroke case mix. Moreover, associations with incident and recurrent vascular events were independent of IL-6 and hsCRP, suggesting that TNF-α signaling may modulate risk in atherosclerosis through a pathologically distinct pathway.

Because circulating TNF levels were associated with both incident and recurrent cardiovascular events, we next examined whether TNF signaling is localized within plaque microenvironments relevant to rupture and thrombosis. Using bulk and spatial transcriptomics, we show that TNF-axis expression is higher in unstable carotid plaque segments and that TNF is more frequently detected at the fibrous cap, where macrophages and CD8+ effector T cells account for most TNF-positive cells. This spatial distribution aligns with histopathological evidence that CD4^+^ and CD8^+^ T cells accumulate in the shoulder region, fibrous cap, and intima of human atherosclerotic lesions.^30^ CD4^+^ T cells recognizing ApoB-100 accumulate in the adventitia surrounding atherosclerotic plaques,^30^ while CD8^+^ effector memory cells clonally expand within plaques and display viral specificity with self-antigen cross-reactivity.^31, 32^ Depletion of CD8^+^ T cells attenuated atherogenesis in aged mice and suggest that these cells actively promote disease.^33^ The identity of the cognate antigens remains uncertain, but emerging evidence suggests that plaque T cells with viral specificities may be reacting to self-antigens with similar homologies to viral epitopes through molecular mimicry, a mechanism implicated in several autoimmune conditions.^31, 32^

Atherosclerotic stroke carries a high residual risk of both recurrent stroke and MACE, with significantly higher recurrence rates than non-atherosclerotic stroke subtypes.^34^ Our work supports evaluation of TNF-related pathways as therapeutic targets for plaque stabilization and vascular prevention after stroke. Whilst there are recognised limitations to prescribing TNF inhibitors (e.g. requirement for exclusion of latent Tuberculosis) and monitoring requirements after commencement (for immunosuppressive effects and malignancy), they are generally well-tolerated in patients deemed appropriate for initiation of therapy. Their safety in older adults has been interrogated and there does not appear to be any increased risk of serious infection or malignancy compared to placebo-matched controls.^35^ Nevertheless, it should be acknowledged that targeting a single node within a complex signaling pathway such as TNF/TNFSF may present challenges and unanticipated off-target effects. For instance, previous RCTs of TNF-inhibitors in patients with heart failure identified an unexpected safety signal with higher rates of decompensated heart failure.^36, 37^ These data emphasise the potential importance of patient selection, disease stage, and timing of treatment initiation of TNF inhibition for prevention. Further mechanistic and early-phase studies are needed to determine safety, and whether systemic TNF-inhibition or more targeted inhibition of plaque-resident TNF-producing immune cells could reduce recurrent vascular events after atherosclerotic stroke and inform targeted selection of patients.

### Strengths and Limitations

We acknowledge limitations. Both the population-based incident stroke analyses and the stroke cohort analyses were observational in design, limiting causal inference and residual confounding cannot be excluded. Therefore, causal effects of TNF signaling for incident and recurrent vascular events can only be established through RCTs of TNF inhibition. Associations between TNF and recurrent stroke may also partly reflect reverse causation, as there is some evidence that brain infarction could influence circulating TNF levels^38^ and timing of measurement varied across studies. However, findings were consistent across studies with diverse populations ranging from TIA-only cohorts to hyperacute disabling stroke. It is also increasingly recognised that interactions between the immune response to brain infarction and atherosclerotic plaque destabilisation are bi-directional. Stroke induces changes to innate immune memory driving chronic organ dysfunction and secondary injury well beyond the acute phase.^39, 40^ Our study was not designed to fully account for the source of the immune stimuli, but rather to interrogate for epidemiological associations between TNF signaling and incident and recurrent vascular events. Future studies with sufficiently large sample sizes should incorporate serial measurements over time of TNF-α, TNFR1, TNFR2, and the TNSF in the days and weeks following acute stroke, to robustly account for dynamic changes in these proteins, and whether these alterations impact on observed associations with recurrent cardiovascular events. The finding that 21 of 34 TNFSF members were independently associated with cardiovascular events may indicate a role for broader inflammatory signal rather than specificity to a single ligands/receptor. Our data does not directly imply that TNF-α inhibition alone will be broadly and consistently protective across the spectrum of patients with established cardiovascular disease. Circulating protein concentrations in UK Biobank were measured using the Olink proximity extension assay and reflect relative rather than absolute protein abundance. Outcomes in the UK Biobank did not permit classification of ischemic stroke subtypes preventing attribution to atherosclerotic pathways. In the IPD cohorts, the low I^2^ zeros shown in Figure 4 may reflect insufficient power to detect true heterogeneity as study cohorts differed in population, timing of phlebotomy and event rate, and thus should be interpreted with a degree of caution. Spatial transcriptomics may miss low-abundance TNF transcripts, and mRNA expression does not necessarily reflect TNF protein abundance or activity. Spatial transcriptomic analyses were also limited to 12 endarterectomy specimens and a targeted gene panel which limits statistical power and molecular coverage. The absence of a robust supportive evidence from Mendelian randomization analysis limits causal inference.

Our work has several strengths. We included data from a very large population-based cohort with long-term follow-up, allowing evaluation of incident events. Access to individual participant–level data enabled replication of findings in independent stroke cohorts with recurrent events, extending evidence to a clinically high-risk population. Observational associations were adjusted for established cardiovascular risk factors and IL-6/CRP, suggesting that the prognostic role of TNF signaling is, at least in part, independent of these factors. Integration with transcriptomic data from human atherosclerotic plaques provides complementary mechanistic support and provides novel insights into the spatial distribution of TNF expression within plaque.

## Conclusion

Circulating TNF pathway proteins were associated with incident stroke and recurrent atherosclerotic cardiovascular events after stroke. In human carotid plaques, TNF-axis expression was higher in unstable segments, while TNF was more frequently detected at the fibrous cap. Together, these convergent findings across population proteomics, secondary prevention cohorts, and plaque biology support further investigation of TNF-directed therapies in atherosclerotic cardiovascular disease.

## Supporting information

PRISMA

## Data Availability

Individual-level UK Biobank data are available to eligible researchers through the UK Biobank application process. This study was conducted under UK Biobank application number 1282644. Access to the individual participant data meta-analysis of the 4 stroke cohorts is available on reasonable request.

## Abbreviations

MACE: Major Adverse Cardiovascular Events
TNF: Tumor Necrosis Factor
TNFR: Tumor Necrosis Factor Receptor
TNFSF: Tumor Necrosis Factor SuperFamily
CAD: Coronary Artery Disease
IL: Interleukin
IPD: Individual Participant Data
PVD: Peripheral Vascular Disease
MI: Myocardial Infarction
CRP: C-Reactive Protein
TIA: Transient Ischemic Attack
RCT: Randomised Controlled Trial

