## Supplemental Methods for "The Role of Tumor Necrosis Factor Signaling in Atherosclerosis and Stroke"

### Study Populations

#### UK Biobank and plasma proteomics

The UK Biobank is a prospective population-based cohort of approximately 500,000 adults aged 40 to 69 years that were recruited between 2006 and 2010 across England, Scotland and Wales.[1] Each participant completed touchscreen questionnaires on sociodemographic and health-related characteristics at their baseline assessment visit. They also underwent physical measurements and provided blood, urine and saliva samples. Participants are followed through linkage to national hospital inpatient records (Hospital Episode Statistics in England, the Scottish Morbidity Record and the Patient Episode Database for Wales) and national death registries. Diagnoses are coded using the International Classification of Diseases, Tenth Revision (ICD-10). UK Biobank derives algorithmically defined outcomes for myocardial infarction and stroke, including ischemic stroke, by combining self-report at baseline with hospital admission and death registry records. It also provides first occurrence fields that record the earliest date of each three-character ICD-10 code across primary care, hospital inpatient, death registry and self-reported sources.

Plasma proteomic profiling was performed by the UK Biobank Pharma Proteomics Project (UKB-PPP).[2] Proteins were measured in 54,219 participants; 46,595 randomly selected from the baseline cohort, 6,376 selected by consortium members and 1,268 who took part in the COVID-19 repeat-imaging study. Proteins were measured with the antibody-based Olink Explore 3072 proximity extension assay, which quantifies 2,941 protein analytes which correspond to 2,923 unique proteins. Protein levels are reported as Normalized Protein eXpression (NPX) values, a relative quantification unit on a $\log_2$ scale.

#### Bulk transcriptomic analysis of human carotid plaques

We used bulk transcriptomic data from the Maastricht Human Plaque Study (MaasHPS; GEO accession GSE163154) that included 43 carotid plaque segments from 24 symptomatic patients undergoing carotid endarterectomy.[3] Endarterectomy specimens were divided into 5mm transverse segments, with alternating segments used for transcriptomic and histological analyses. Plaques were classified by two pathologists using a modified American Heart Association classification. Stable segments (n=16) included pathological intimal thickening, early or late fibroatheroma and fibrocalcified plaques, while unstable segments (n=27) included intraplaque haemorrhage, plaque rupture or acute occlusion. Intraplaque haemorrhage was assessed by quantifying extravascular erythrocyte deposits on adjacent haematoxylin and eosin-stained sections. Gene expression was measured using the Illumina HumanRef-8 v2 microarray.

**Spatial transcriptomic dataset of human carotid plaques**

Spatial transcriptomic data were obtained from a published dataset of human carotid plaques (GEO accession GSE294466).[4] Carotid endarterectomy specimens (12 advanced plaques and 4 paired early lesions) from 12 patients with carotid stenosis were obtained from the Munich Vascular Biobank. Formalin-fixed paraffin-embedded sections were profiled on the 10x Genomics Xenium platform using two custom panels that included 548 unique genes. Cell segmentation was performed with Baysor, and cell types were annotated manually by clustering and differential expression. A cardiovascular pathologist annotated histological subregions (media, intima, necrotic core, fibrous cap and lumen) on hematoxylin and eosin stains.

**Eligibility criteria for Systematic Review**

Inclusion criteria:

Adults with history of Ischaemic Stroke/TIA with TNF-Alpha measured in blood after the index event, and available reported associations on recurrent vascular events (stroke, major adverse cardiac events).

Exclusion criteria:

1. Age <18 years
2. Non-human studies.
3. Studies related to Haemorrhagic Stroke.
4. Incomplete/unavailable outcome data.
5. Incomplete/unavailable timed TNF-Alpha measurement or measurements taken before the index stroke.
6. Duplicate studies, crossover studies, cross-sectional studies, editorials, case reports, letters, review articles and expert opinion articles.

**Search Strategy for Systematic Review**

Records were screened by an individual reviewer for title and abstract, and full text screening performed by two reviewers JB and PS. A third reviewer JMcC was available to resolve disagreements between the two reviewers. Previously unpublished data were considered and included if eligibility criteria were met. Full search strategy and inclusion/exclusion criteria available below.

<u>Search Terms</u>

OVID Medline

1. exp Stroke/
2. exp Brain Ischemia/
3. Brain isch?emia.mp
4. cerebrovascular accident*.mp
5. isch?emic stroke.mp
6. (Brain adj1 (isch?emi* or infarct* or embol* or thromb*)).tw.
7. (cereb* adj1 (isch?emi* or infarct* or embol* or thromb*)).tw.
8. Cortical infarct*.mp
9. exp Carotid Stenosis/
10. Carotid stenosis.mp
11. exp Carotid Artery Thrombosis/
12. Carotid artery thrombosis.mp
13. exp Carotid Artery Diseases/
14. carotid artery disease.mp
15. internal carotid artery occlusion.mp
16. intracranial atherosclerosis.mp. or exp Intracranial Arteriosclerosis/
17. exp Intracranial Embolism/
18. exp Intracranial Thrombosis/
19. exp “Intracranial Embolism and Thrombosis"/
20. cerebral artery occlusion.mp
21. intracranial occlusion.mp
22. large vessel occlusion.mp
23. exp Ischemic Attack, Transient/
24. tia.mp25. transient isch?emi* attack.mp
26. transient isch?emi* event.mp
27. cardioembolic stroke.mp

28. cardio-embolic stroke.mp
29. Lacunar stroke.mp
30. exp Vertebrobasilar Insufficiency/
31. 1 or 2 or 3 or 4 or 5 or 6 or 7 or 8 or 9 or 10 or 11 or 12 or 13 or 14 or 15 or 16 or 17 or 18 or 19 or 20 or 21 or 22 or 23 or 24 or 25 or 26 or 27 or 28 or 29 or 30
32. exp Tumor Necrosis Factor- alpha/
33. tumo?r necrosis factor al* .tw.
34. tnf al* .mp
35. cachectin.mp
36. endotoxin induced factor.mp
37. endo* induc* factor.tw.
38. Differentiation inducing factor.mp
39. Diff* induc* factor.mp
40. 32 or 33 or 34 or 35 or 36 or 37 or 38 or 39
41. exp Recurrence/
42. recurrent disease.mp
43. recur*.tw
44. (recur* adj3 stroke).tw
45. (recur* adj3 event).mp
46. exp Outcome Assessment, Health Care/
47. predictive value.mp
48. exp Secondary Prevention/
49. Secondary prevention.mp
50. exp Prognosis/
51. exp "Cause of Death"/
52. exp Death, Sudden, Cardiac/
53. (vascular adj3 death*).mp
54. (vascular adj3 mortality)).tw.
55. (cardiovascular adj2 (death* or event*)).tw.
56. (cardiac adj2 (death* or event*)).tw.
57. 41 or 42 or 43 or 44 or 45 or 46 or 47 or 48 or 49 or 50 or 51 or 52 or 53 or 54 or 55 or 56
58. 31 and 40 and 57
59. limit 58 to (english language and humans)

60. limit 59 to abstracts

EMBASE

1. 'cerebrovascular accident'/exp OR 'cerebrovascular accident'
2. stroke:ti,ab
3. 'isch?mic stroke':ti,ab
4. brain NEAR/3 isch?mi*
5. isch?mi* NEAR/3 cereb*
6. 'carotid artery thrombosis'/exp
7. 'carotid artery disease'/exp
8. 'carotid stenosis':ti,ab
9. 'internal carotid artery occlusion':ti,ab
10. 'intracranial atherosclerosis'/exp
11. 'intracranial atherosclerosis':ti,ab
12. 'cerebral artery occlusion'/exp
13. 'cerebral artery occlusion':ti,ab
14. 'large vessel occlusion'/exp
15. 'large vessel occlusion':ti,ab
16. 'intracranial thrombosis'/exp
17. 'intracranial thrombosis':ti,ab
18. 'intracranial occlusion':ti,ab
19. 'intracranial embolism'/de
20. 'intracranial embolism':ti,ab
21. 'tia'/de
22. 'transient isch?mi* attack':ti,ab
23. 'transient isch?mi* event':ti,ab
24. 'cardioembolic stroke'/exp
25. 'lacunar stroke'/exp
26. 'cortical infarct*':ti,ab
27. 'cereb* infarct*':ti,ab
28. 'brain infarct*':ti,ab
29. 'wallenberg* syndrome'
30. 'posterior circulation stroke':ti,ab
31. #1 OR #2 OR #3 OR #4 OR #5 OR #6 OR #7 OR #8 OR #9 OR #10 OR #11 OR #12

OR #13 OR #14 OR #15 OR #16 OR #17 OR #18 OR #19 OR #20 OR #21 OR #22 OR #23 OR #24 OR #25 OR #26 OR #27 OR #28 OR #29 OR #30

32. 'tumor necrosis factor'/exp
33. 'serum tum*r necrosis factor':ti,ab
34. 'blood tum*r necrosis factor':ti,ab
35. 'tnf al*a':ti,ab
36. 'cachectin':ti,ab
37. 'endotoxin induced factor'
38. 'differentiation inducing factor'/exp
39. #32 OR #33 OR #34 OR #35 OR #36 OR #37 OR #38
40. 'recurrent disease'/exp
41. 'recur*':ti,ab
42. recur* NEAR/3 stroke
43. recur* NEAR/4 event44. 'outcome assessment'/exp
45. 'predictive value'/exp
46. 'predictive value':ti,ab
47. 'secondary prevention'/exp
48. 'secondary prevention':ti,ab
49. 'prognosis'/exp
50. 'cause of death'/de
51. 'sudden cardiac death'/de
52. vascular NEAR/3 death*
53. cardiovascular NEAR/2 event
54. 'vascular death*'
55. cardiovascular NEAR/3 death
56. #40 OR #41 OR #42 OR #43 OR #44 OR #45 OR #46 OR #47 OR #48 OR #49 OR #50 OR #51 OR #52 OR #53 OR #54 OR #55
57. #31 AND #39 AND #56
58. #31 AND #39 AND #56 AND [humans]/lim AND [english]/lim AND [abstracts]/lim AND [embase]/lim

**Collection and Harmonisation of IPD**

Authors of eligible studies were invited to provide IPD for analysis. 3 studies provided IPD for centralized analysis. One study group (PROGRESS) provided pre-specified identical remote analysis and provided summary aggregate data for meta-analysis.

**IPD Collected from Participating Studies**

Key individual participant data requested from collaborating sites included:

Demographics

- Age
- Gender
- Ethnicity

Index event

- Date of event
- Subtype:
    - TIA (cerebral or retinal) or ischemic stroke (cerebral infarction or retinal event)
    - TIA was defined as a focal neurological event lasting <24hours due to an ischemic cause for all studies
- National Institute for Health Stroke Scale (NIHSS). Where this was not available, the Scandinavian Stroke Scale (SSS) was used to derive an estimated NIHSS using the following equation: NIHSS=2 5.68 – 0.43 * SSS. This formula has been shown to approximate the NIHSS with good accuracy.
- Classification of stroke mechanism in the opinion of the study investigators

Cardiovascular risk factors:

- Hypertension, defined as:
    - A new, self-reported or documented history of hypertension, or
    - Use of blood pressure medications prior to the index event
- Hyperlipidaemia defined as:
    - Documented or self-reported history of hypercholesterolaemia, or
    - Use of lipid lowering agents prior to the index event (eg. Statin, ezetimibe, fibrates or proprotein convertase subtilisin/Kexin type 9 [PCSK9] inhibitors), or
    - A new diagnosis of hyperlipidaemia based on Adult Treatment Panel III (ATP III) criteria.
- Diabetes mellitus, defined as:

    - A new, self-reported or documented history of diabetes mellitus, or
    - Use of glucose lowering mediations prior to the index event
  - Coronary artery disease, defined as any pre-existing history of angina, acute coronary syndrome or coronary revascularisation procedure
  - Atrial fibrillation, defined as any new, self-reported or documented history of permanent or paroxysmal atrial fibrillation
  - Smoking status: current/ex-smoker/never smoker.

Medications around the time of enrolment

  - Statin therapy pre-event or at the end of the treatment episode for the index event
  - Anti-thrombotic therapy pre-event or at the end of the treatment episode for the index event. This included anti-coagulants (apixaban, dabigatran, edoxaban, rivaroxaban, warfarin) and/or anti-platelets (aspirin, clopidogrel, ticagrelor, prasugrel, dipyridamole, cilostazol).

Blood inflammatory markers:

- First available measurement of Tumour Necrosis Factor Alpha after the index event
- Time from index event to phlebotomy

Follow up (inclusive of vascular recurrence events during follow up)

- Date of last follow-up
- First recurrent fatal or non-fatal stroke (as defined by the study) during follow up and date of first recurrence (if applicable)
- First recurrent fatal or non-fatal cardiac event (as defined by the study) during follow up and date of first recurrence (if applicable)

Key study level data requested

- Eligibility criteria
- Participating sites
- Screening and recruitment for enrolment
- Procedure (if present) for screening for infection or co-morbid inflammatory conditions
- Sample type, storage conditions, assay used and method of detection of TNF Alpha
- Outcome definitions and outcome adjudication processes
- Procedures for follow up

**Risk of bias assessment of included studies**

The Quality in Prognostic Studies (QUIPS) tool was adjudicated by 2 reviewers to assess the risk of bias of included studies. QUIPS evaluates six domains of potential bias in prognostic research: (i) study participation; (ii) study attrition; (iii) prognostic factor measurement; (iv) confounder measurement; (v) outcome measurement; (vi) analysis/reporting. As the analysis/reporting domain was standardized across participating studies in this analysis, this domain was not assessed.

**Definition of outcomes analysed in Meta Analysis**

The pre-specified co-primary outcomes were:

1. Recurrent MACE (composite of stroke, MI, unstable angina, cardiac arrest, cardiac or vascular death)
2. Recurrent stroke (ischaemic, haemorrhagic, or unspecified) occurring after blood sampling.

Recurrences that occurred prior to blood sampling of TNF-Alpha were excluded.

**Description of Included Studies in IPD**

The BICHUS (Biobanco de Ictus del Complejo Hospitalario Universitario de Santiago de Compostela registry) was a prospective observational study of consecutive patients with acute ischemic stroke admitted to the Stroke Unit of the University Clinical Hospital of Santiago de Compostela, Spain from September 2007 to March 2018.[26] Patients were eligible for this analysis if they had measurements for a blood inflammatory marker.

Exclusion criteria were primary intracerebral haemorrhage, or presence of an acute or chronic inflammatory process. All patients were followed up in person every six hours during hospitalisation and three months after hospital discharge. Suspected recurrent events were evaluated by a trained neurologist blinded to baseline inflammatory marker levels.

REGITELL (Registro de pacientescon AIT de Lleida) was a single-centred prospective cohort observational study of consecutive patients with diagnosed TIA attending the emergency department of Hospital Universitari Arnau de Vilanova de Lleida, Spain between January 2008 and January 2012.[28] Patients with ischemic stroke, TIA mimics, pre-existing disability (mRS >3), advanced chronic kidney disease, infection or inflammatory disorders were excluded. Suspected recurrences were evaluated via clinical interviews performed during follow-up.

BIOSTROKE-TIA (Biomarkers and Imaging of ischemic Stroke or TIA) was a prospective multi-centred observational cohort study of patients with recent (<14 days) non-severe (mRS ≤3) ischemic stroke or TIA (<48 hours), attending the emergency department or rapid-access TIA clinics of five stroke centres in Dublin, Ireland, between 2008-2012.[29] Key exclusion criteria were: asymptomatic stroke on brain imaging, infection or pro-inflammatory co-morbidities. All patients were followed up in-person or by telephone at seven, 28, 90 days and one year by trained study staff and assessed for recurrence of stroke, disability, and other vascular events. Outcomes were centrally adjudicated by a single investigator, blinded to baseline clinical data and inflammatory markers.

PROGRESS (Perindopril Protection Against Recurrent Stroke Study) was a randomised double-blind placebo controlled trial of 6105 individuals from 172 centres in Asia, Australasia, and Europe.[27] Patients were randomly assigned active treatment (n=3051) or placebo (n=3054). Eligible patients had a history of stroke (intracerebral haemorrhage or ischaemic stroke) or TIA within the previous 5 years (but more than 1 month prior).

Participants were required to have no definite indication (such as heart failure) for treatment with an angiotensin-converting-enzyme (ACE) inhibitor and no definite contraindication (such as previous intolerance) to such treatment. Active treatment comprised a flexible regimen based on perindopril (4 mg daily) with the addition of indapamide (2·5 mg daily). In the year after randomisation, participants were seen on five occasions. In the second and subsequent years, visits were held every 6 months. An endpoint adjudication committee reviewed source documentation for all individuals who had a suspected stroke or myocardial infarction or who died during the scheduled period of follow-up. For the purposes of this analysis, a nested case-control study was performed and patients whose qualifying event was haemorrhagic stroke were excluded. Cases were defined as those with recurrent stroke or MACE during follow-up. Using standard methodology for nested case-control studies, each case was randomly matched to between 1 and 3 controls selected from all those free of recurrent stroke or MACE. Matching variables were age (within 5 years), sex, treatment allocated (active/placebo), therapy (mono/dual), region (Australia or New Zealand/China/Japan/France or Belgium/Italy/Sweden/UK or Ireland) and most recent qualifying event.
