## Supplementary material for "The Role of Tumor Necrosis Factor Signaling in Atherosclerosis and Stroke": Figure S

**Supplementary Figure 1**. Cumulative incidence of ischemic stroke by third of each circulating TNF superfamily protein.

Unadjusted cumulative incidence by third of plate-adjusted protein level. P values are from log-rank tests across thirds. Follow-up was censored at death or end of follow-up and is shown to 15 years. T1 indicates lowest third; T3, highest third.

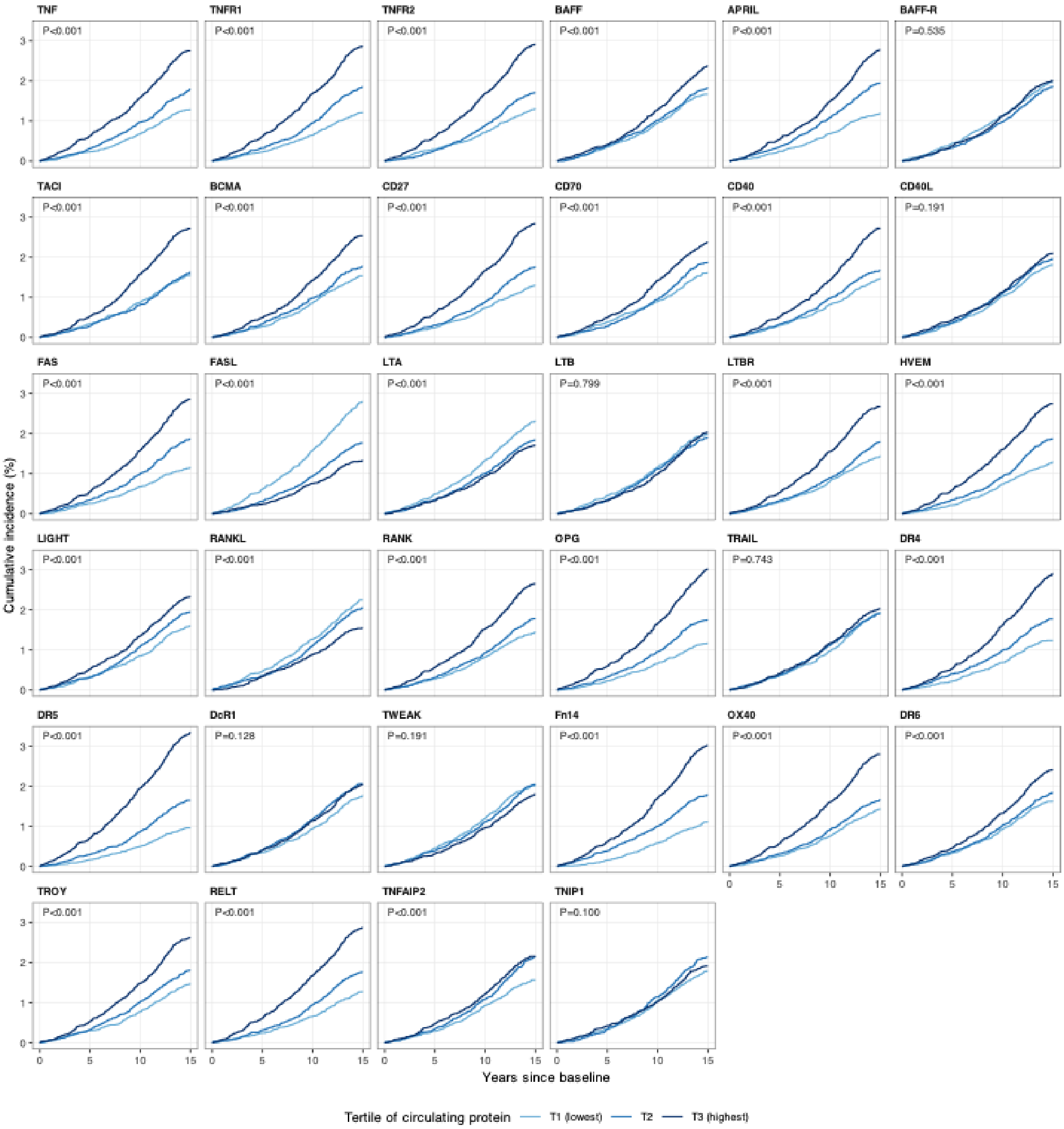

**Supplementary Figure 2**. Cumulative incidence of myocardial infarction by third of each circulating TNF superfamily protein.

Unadjusted cumulative incidence by third of plate-adjusted protein level. P values are from log-rank tests across thirds. Follow-up was censored at death or end of follow-up and is shown to 15 years. T1 indicates lowest third; T3, highest third.

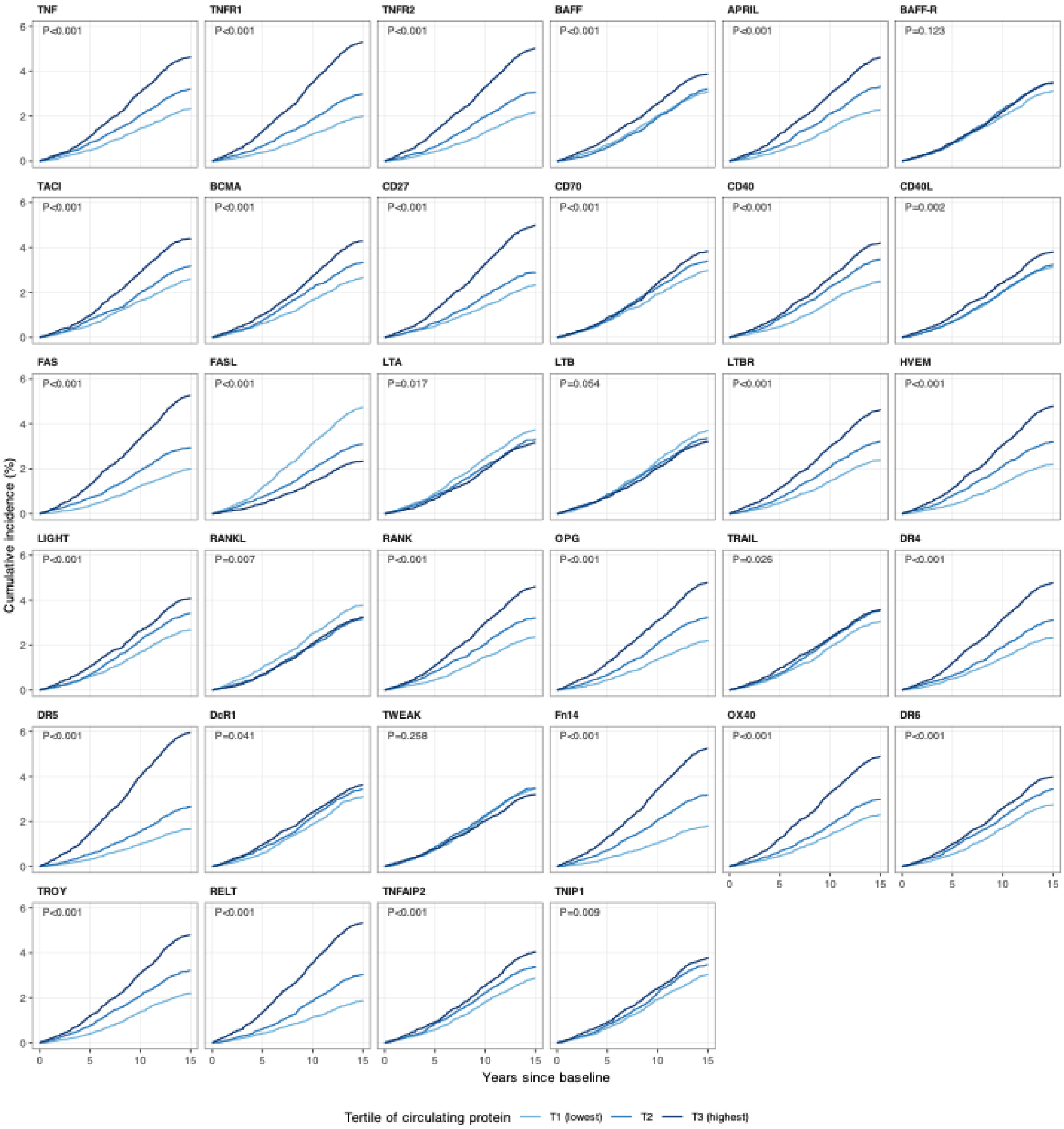

**Supplementary Figure 3**. Cumulative incidence of peripheral vascular disease by third of each circulating TNF superfamily protein.

Unadjusted cumulative incidence by third of plate-adjusted protein level. P values are from log-rank tests across thirds. Follow-up was censored at death or end of follow-up and is shown to 15 years. T1 indicates lowest third; T3, highest third.

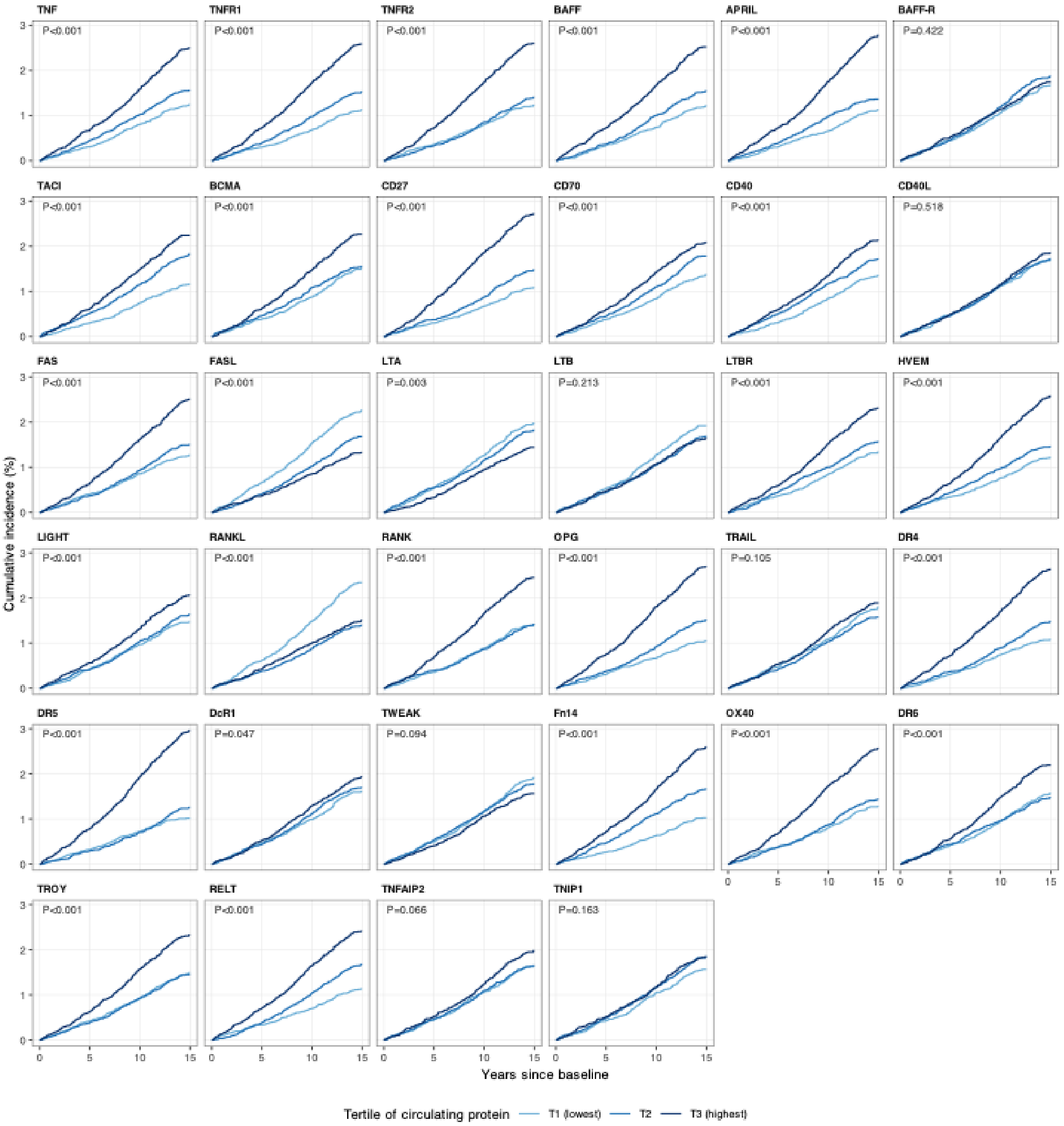

**Supplementary Figure 4.** TNF Superfamily proteins and risk of incident cardiovascular disease after adjustment for IL6 and CRP.

Hazard ratios per SD increase in circulating protein levels. Blue circles: adjusted for age, sex, ethnicity, Townsend deprivation index, education, smoking, alcohol, BMI, systolic blood pressure, and diabetes. Red triangles: additionally adjusted for log-CRP and IL-6. P-values are FDR-adjusted for multiple comparisons.

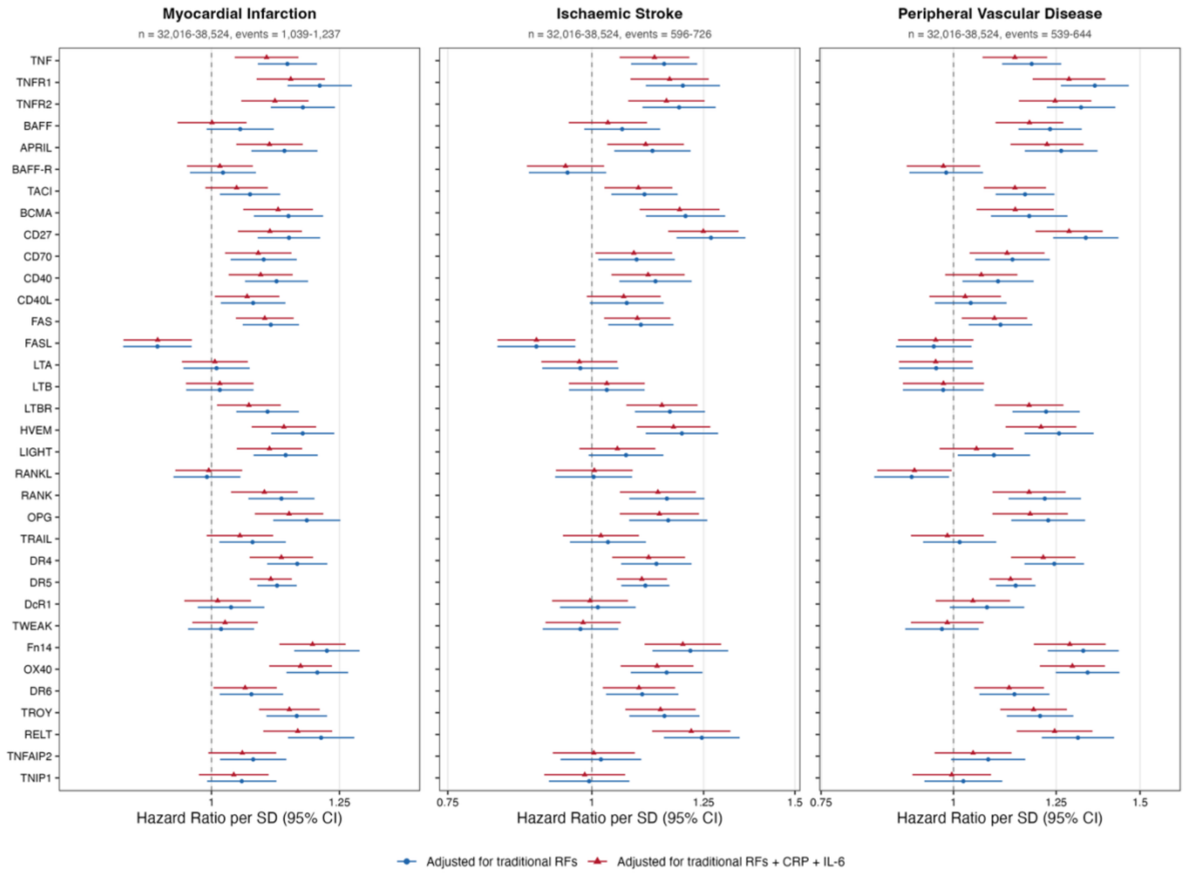

**Supplementary Figure 5.** TNF Superfamily Proteins and risk of incident cardiovascular disease in 2-year landmark analysis.

Hazard ratios per SD increase in circulating protein levels. Both analyses adjusted for age, sex, ethnicity, Townsend deprivation index, education, smoking, alcohol, BMI, systolic blood pressure, and diabetes. Blue circles: primary analysis. Red triangles: 2-year landmark analysis (excluding events in first 2 years of follow-up).

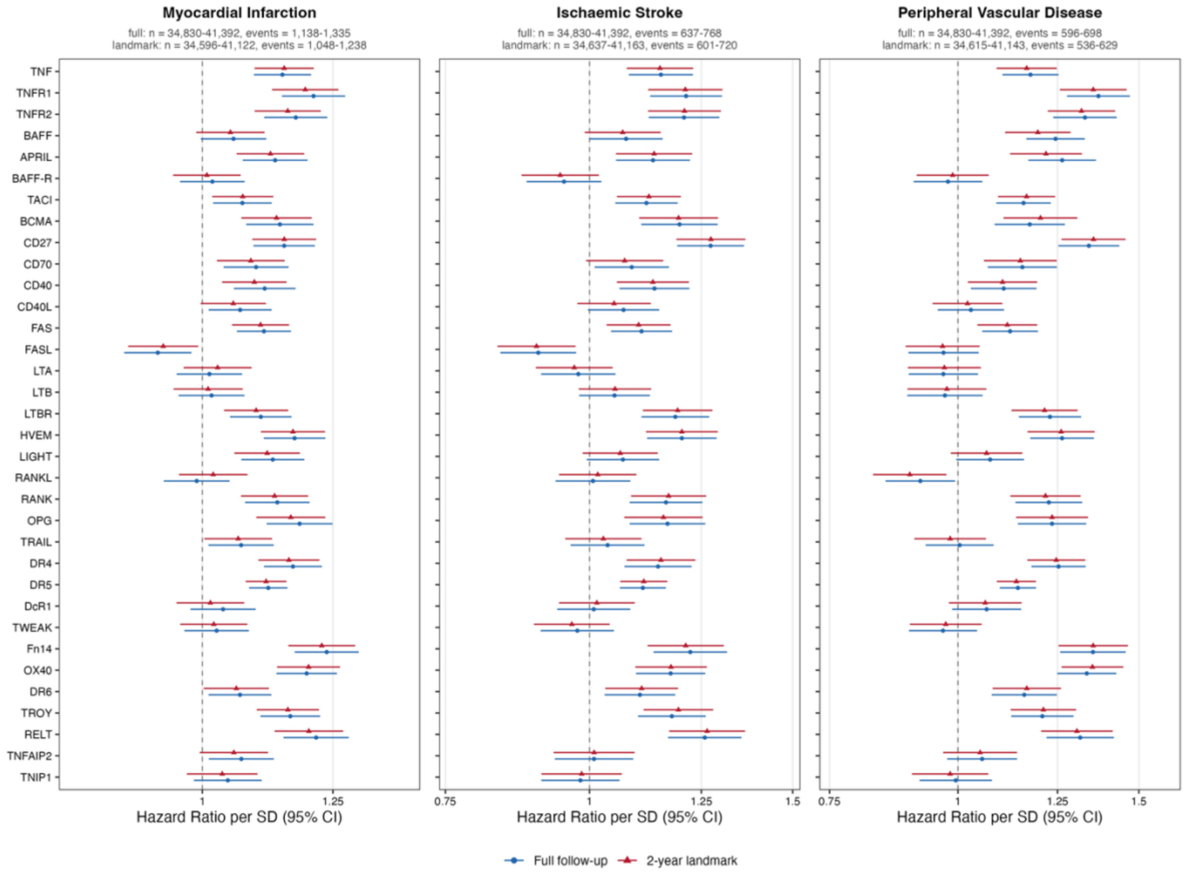

**Supplementary Figure 6.** TNF Superfamily proteins and risk of incident cardiovascular disease, excluding participants with CRP >10 mg/L at baseline

Hazard ratios per SD increase in circulating protein levels. Both analyses adjusted for age, sex, ethnicity, Townsend deprivation index, education, smoking, alcohol, BMI, systolic blood pressure, and diabetes. Blue circles: primary analysis. Red triangles: excluding participants with baseline CRP >10 mg/L at baseline.

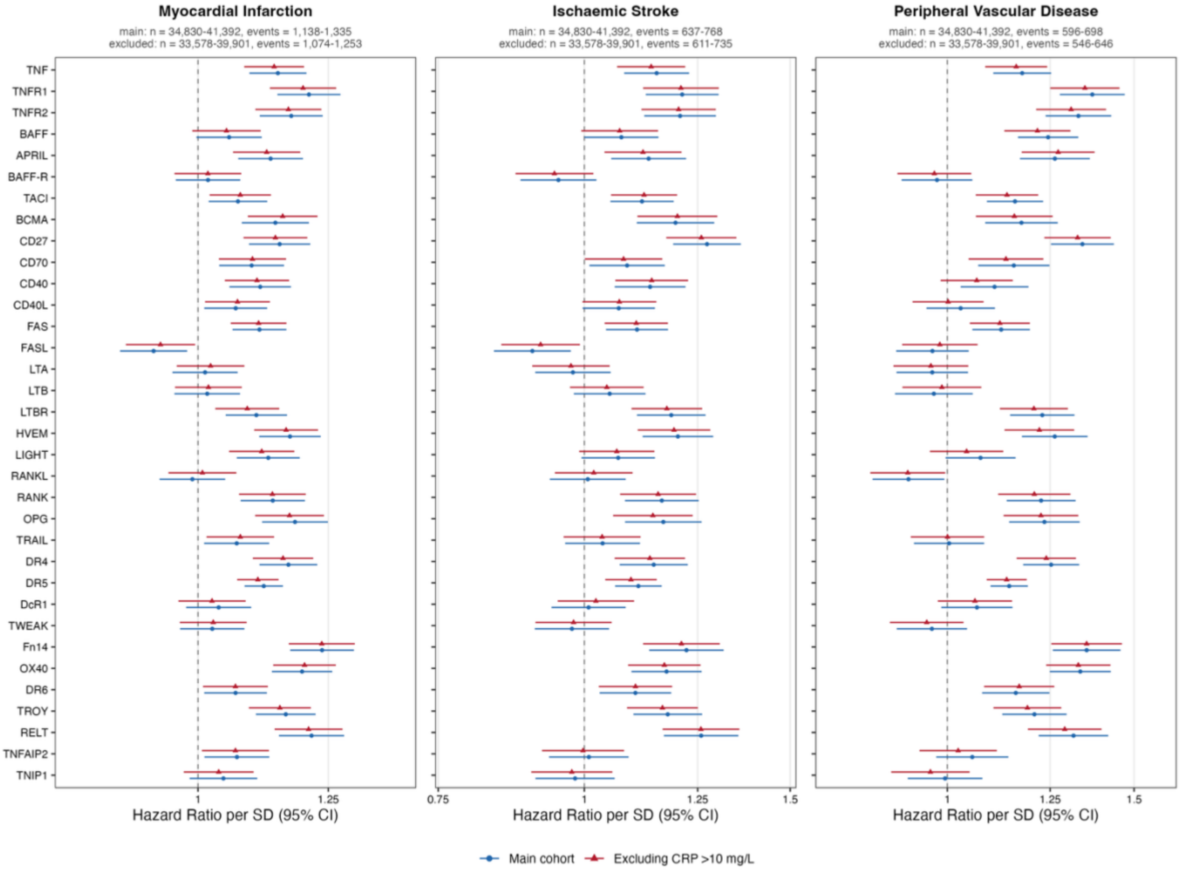

**Supplementary Figure 7.** TNF Superfamily proteins and risk of incident cardiovascular disease, by period of follow-up

Hazard ratios per SD increase in circulating protein levels. Both analyses adjusted for age, sex, ethnicity, Townsend deprivation index, education, smoking, alcohol, BMI, systolic blood pressure, and diabetes. Blue circles: 0–5 years of follow-up. Gray squares: 5–10 years. Red triangles: >10 years. Each period includes participants event-free at its start, with follow-up censored at its end.

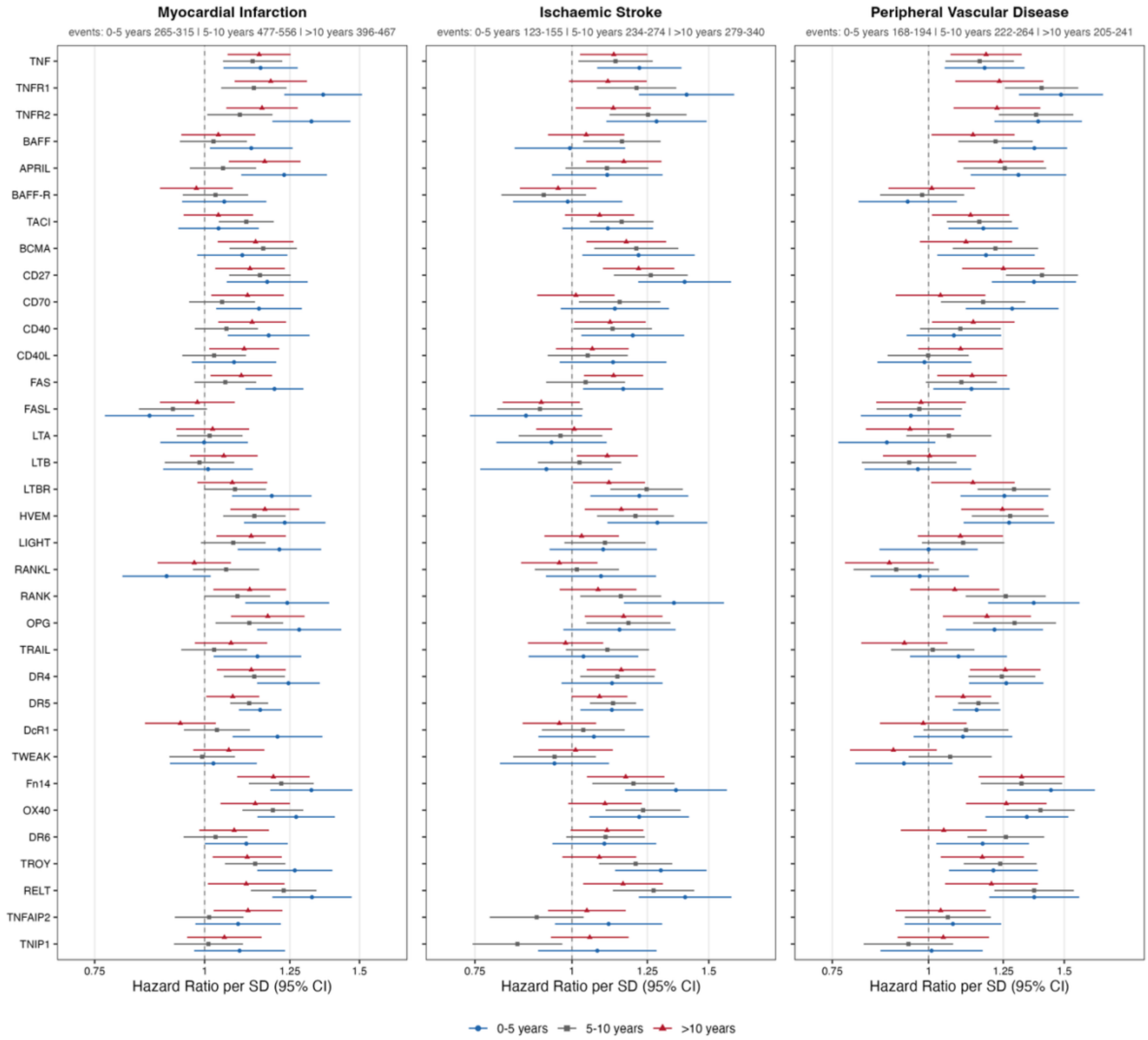

**Supplementary Figure 8. PRISMA (Preferred Reporting Items for Systematic Reviews and Meta-Analyses) flow-chart for study inclusion.**

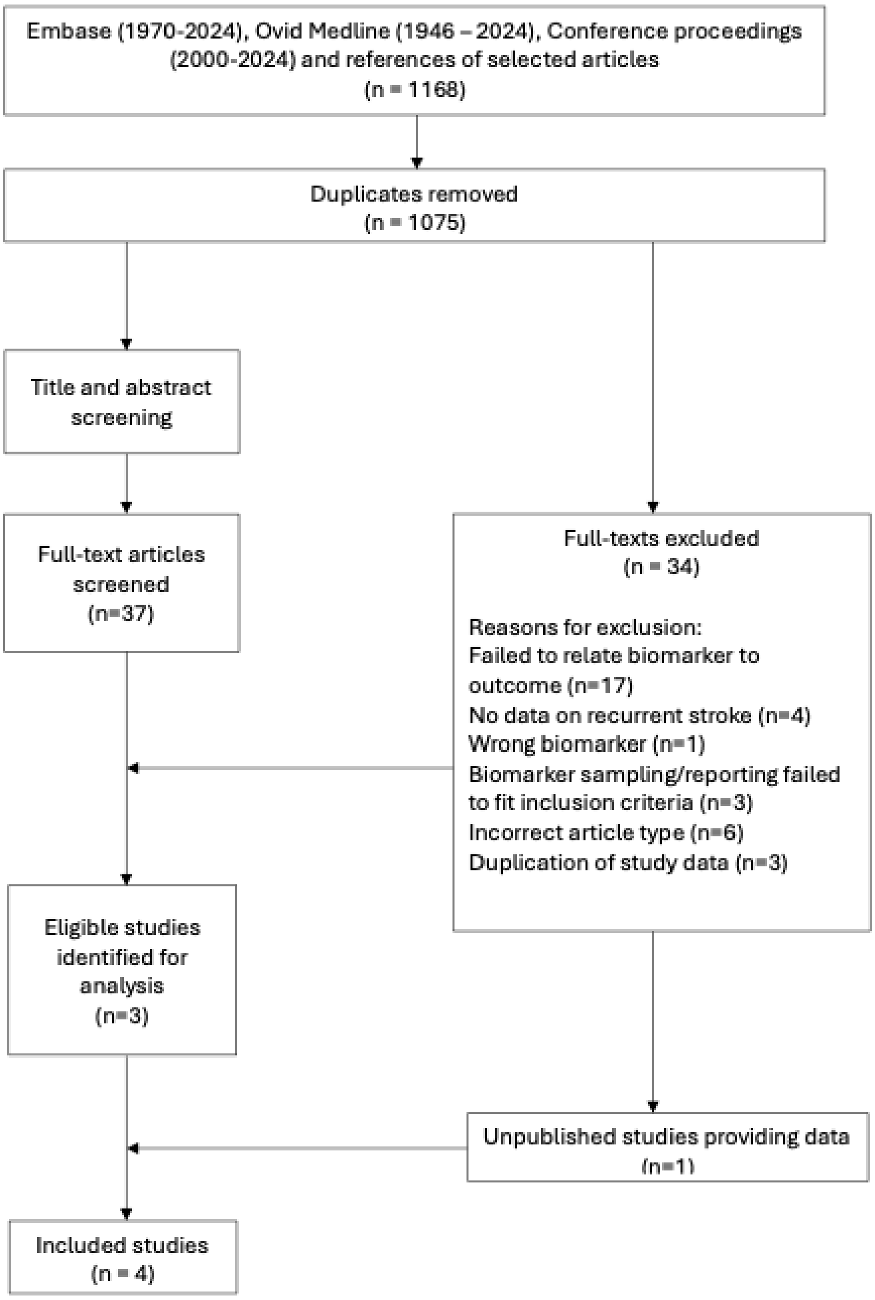

**Supplementary Figure 9**: Leave one out sensitivity analysis performed for (a) TNF alpha and recurrent stroke and (b) TNF alpha and recurrent MACE. Analysis was performed per log unit increase in TNF alpha and adjusted for age, sex, index event, hypertension, smoking, diabetes mellitus, coronary artery disease, atrial fibrillation, statin and antithrombotic use and trial arm (where appropriate).

**Figure 9a**: Association of TNF alpha and recurrent stroke; Leave One Out Analysis per $\log_e$-unit increase in TNF alpha:

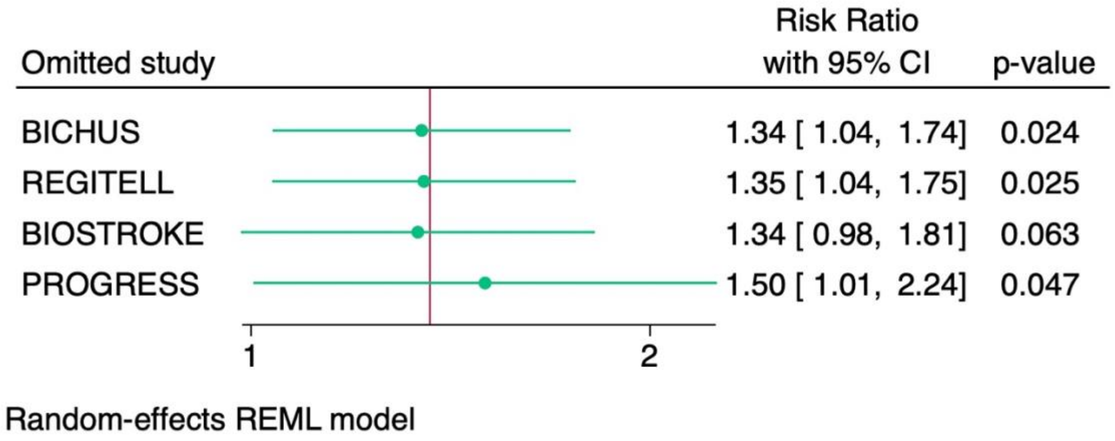


**Figure 9b:** Association of TNF alpha and recurrent MACE; Leave One Out Analysis per $\log_e$-unit increase in TNF alpha.

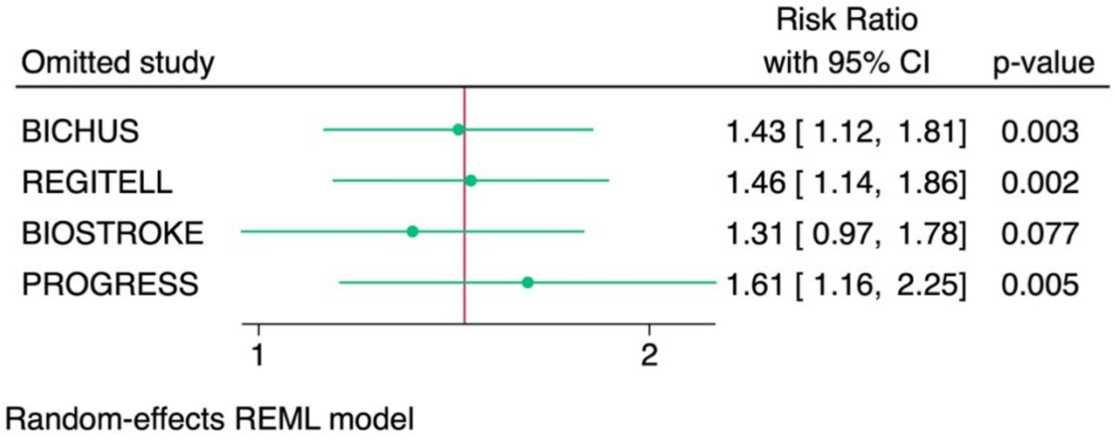
