## Supplementary material for "The Role of Tumor Necrosis Factor Signaling in Atherosclerosis and Stroke": PRISMA

| PRISMA-IPD Section/topic | Item No | Checklist item | Reported on page |
|---|---|---|---|
| **Title** | 1 | Identify the report as a systematic review and meta-analysis of individual participant data. | 4 |
| Title | | | |
| **Abstract** | 2 | | |
| Structured summary | | Provide a structured summary including as applicable: | 4 |
| | | **Background**: state research question and main objectives with information on participants, interventions, comparators and outcomes. | 4 |
| | | **Methods**: report eligibility criteria; data sources including dates of last bibliographic search or elicitation, noting that IPD were sought; methods of assessing risk of bias. | 4 |
| | | **Results:** provide number and type of studies and participants identified and number (%) obtained; summary effect estimates for main outcomes (benefits and harms) with confidence intervals and measures of statistical heterogeneity. Describe the direction and size of summary effects in terms meaningful to those who would put findings into practice. | 4-5 |
| | | Discussion: state main strengths and limitations of the evidence, general interpretation of the results and any important implications. | 5 |
| | | Other: report primary funding source, registration number and registry name for the systematic review and IPD meta-analysis | N/A |
| **Introduction** | | | |
| Rationale | 3 | Describe the rationale for the review in the context of what is already known. | 6/7 |
| Objectives | 4 | Provide an explicit statement of the questions being addressed with reference, as applicable, to participants, interventions, comparisons, outcomes and study design (PICOS). Include any hypotheses that relate to particular types of participant-level subgroups. | 6 |
| **Methods** | | | |
| Protocol and registration | 5 | Indicate if a protocol exists and where it can be accessed. If available, provide registration information including registration number and registry name. Provide publication details, if applicable. | 8 |
| Eligibility criteria | 6 | Specify inclusion and exclusion criteria including those relating to participants, interventions, comparisons, outcomes, study design and characteristics (e.g. years when conducted, required minimum follow-up). Note whether these were applied at the study or individual level i.e. whether eligible participants were included (and ineligible participants excluded) from a study that included a wider population than specified by the review inclusion criteria. The rationale for criteria should be stated. | 8, Supplement |
| Identifying studies – information sources | 7 | Describe all methods of identifying published and unpublished studies including, as applicable: which bibliographic databases were searched with dates of coverage; details of any hand searching including of conference proceedings; use of study registers and agency or company databases; contact with the original research team and experts in the field; open adverts and surveys. Give the date of last search or elicitation. | 8, Supplement |
| Identifying studies – search | 8 | Present the full electronic search strategy for at least one database, including any limits used, such that it could be repeated. | Supplement |
| Study selection processes | 9 | State the process for determining which studies were eligible for inclusion | Supplement |
| Data collection processes | 10 | Describe how IPD were requested, collected and managed, including any processes for querying and confirming data with investigators. If IPD were not sought from any eligible study, the reason for this should be stated (for each such study). If applicable, describe how any studies for which IPD were not available were dealt with. This should include whether, how and what aggregate data were sought or extracted from study reports and publications (such as extracting data independently in duplicate) and any processes for obtaining and confirming these data with investigators. | 8, Supplement |
| Data items | 11 | Describe how the information and variables to be collected were chosen. List and define all study level and participant level data that were sought, including baseline and follow-up information. If applicable, describe methods of standardising or translating variables within the IPD datasets to ensure common scales or measurements across studies. | Supplement |
| IPD integrity | A1 | Describe what aspects of IPD were subject to data checking (such as sequence generation, data consistency and completeness, baseline imbalance) and how this was done. | Supplement |
| Riks of bias assessment | 12 | Describe methods used to assess risk of bias in the individual studies and whether this was applied separately for each outcome. If applicable, describe how findings of IPD checking were used to inform the assessment. Report if and how risk of bias assessment was used in any data synthesis. | Supplement, STable 9 |
| Specification of outcomes and effect measures | 13 | State all treatment comparisons of interests. State all outcomes addressed and define them in detail. State whether they were pre-specified for the review and, if applicable, whether they were primary/main or | Supplement |

| | | | |
|---|---|---|---|
| | | secondary/additional outcomes. Give the principal measures of effect (such as risk ratio, hazard ratio, difference in means) used for each outcome. | |
| Synthesis methods | 14 | Describe the meta-analysis methods used to synthesise IPD. Specify any statistical methods and models used. Issues should include (but are not restricted to):<br>• Use of a one-stage or two-stage approach.<br>• How effect estimates were generated separately within each study and combined across studies (where applicable).<br>• Specification of one-stage models (where applicable) including how clustering of patients within studies was accounted for.<br>• Use of fixed or random effects models and any other model assumptions, such as proportional hazards.<br>• How (summary) survival curves were generated (where applicable).<br>• Methods for quantifying statistical heterogeneity (such as I2 and t2).<br>• How studies providing IPD and not providing IPD were analysed together (where applicable).<br>• How missing data within the IPD were dealt with (where applicable). | 9 |
| Exploration of variation in effect | A2 | If applicable, describe any methods used to explore variation in effects by study or participant level characteristics (such as estimation of interactions between effect and covariates). State all participant-level characteristics that were analysed as potential effect modifiers, and whether these were pre-specified. | 9 |
| Risk of bias across studies | 15 | Specify any assessment of risk of bias relating to the accumulated body of evidence, including any pertaining to not obtaining IPD for particular studies, outcomes or other variables. | Supplement |
| Additional analyses | 16 | Describe methods of any additional analyses, including sensitivity analyses. State which of these were pre-specified. | 9 |
| **Results** | | | |
| Study selection and IPD obtained | 17 | Give numbers of studies screened, assessed for eligibility, and included in the systematic review with reasons for exclusions at each stage. Indicate the number of studies and participants for which IPD were sought and for which IPD were obtained. For those studies where IPD were not available, give the numbers of studies and participants for which aggregate data were available. Report reasons for non-availability of IPD. Include a flow diagram. | 14, Supplement |
| Study characteristics | 18 | For each study, present information on key study and participant characteristics (such as description of interventions, numbers of participants, demographic data, unavailability of outcomes, funding source, and if applicable duration of follow-up). Provide (main) citations for each study. Where applicable, also report similar study characteristics for any studies not providing IPD. | 14, Table 1, Supplement |
| IPD integrity | A3 | Report any important issues identified in checking IPD or state that there were none. | None identified |
| Riks of bias within studies | 19 | Present data on risk of bias assessments. If applicable, describe whether data checking led to the up-weighting or down-weighting of these assessments. Consider how any potential bias impacts on the robustness of meta-analysis conclusions. | Supplement, STable 9 |
| Results of individual studies | 20 | For each comparison and for each main outcome (benefit or harm), for each individual study report the number of eligible participants for which data were obtained and show simple summary data for each intervention group (including, where applicable, the number of events), effect estimates and confidence intervals. These may be tabulated or included on a forest plot. | Table 2 |
| Results of syntheses | 21 | Present summary effects for each meta-analysis undertaken, including confidence intervals<br>and measures of statistical heterogeneity. State whether the analysis was pre-specified, and report the numbers of studies and participants and, where applicable, the number of events<br>on which it is based. When exploring variation in effects due to patient or study characteristics, present summary interaction estimates for each characteristic examined, including confidence<br>intervals and measures of statistical heterogeneity. State whether the analysis was pre- specified. State whether any interaction is consistent across trials. Provide a description of the direction and size of effect in terms meaningful to those who would put findings into practice. | 14, Table 2, Figure 4 |
| Risk of bias across studies | 22 | Present results of any assessment of risk of bias relating to the accumulated body of evidence, including any pertaining to the availability and representativeness of available studies, outcomes or other variables. | Supplement, STable 9 |
| Additional analyses | 23 | Give results of any additional analyses (eg. Sensitivity analyses). If application, this should also include any analyses that incorporate aggregate data for studies that do not have IPD. If application summarise the main meta-analysis results following the inclusion or exclusion of studies for which IPD were not available. | STables 13-16 |
| **Discussion** | | | |

| | | | |
|---|---|---|---|
| Summary of evidence | 24 | Summarise the main findings, including the strength of evidence for each main outcome. | 19, 20, 21 |
| Strengths and limitations | 25 | Discuss any important strengths and limitations of the evidence including the benefits of access to IPD and any limitations arising from IPD that were not available. | 24, 25 |
| Conclusions | 26 | Provide a general interpretation of the findings in the context of other evidence. | 26 |
| Implications | A4 | Consider relevance to key groups (such as policy makers, service providers and service users). Consider implications for future research. | 23, 26 |
| **Funding** | | | |
| Funding | 27 | Describe sources of funding and other support (such as supply of IPD), and the role in the systematic review of those providing such support | N/A |
